# Community types of the human gut virome are associated with endoscopic outcome in ulcerative colitis

**DOI:** 10.1101/2022.07.17.22277421

**Authors:** Daan Jansen, Gwen Falony, Sara Vieira-Silva, Ceren Simsek, Tine Marcelis, Clara Caenepeel, Kathleen Machiels, Jeroen Raes, Séverine Vermeire, Jelle Matthijnssens

## Abstract

**Objective:** IBD patients have an altered gut virome composition; however, the relationship to disease is unknown. Our aim is to investigate the existence of viral community types and assess the impact of therapeutic outcome (and other covariates) on the gut virome.

**Design:** Viral particle enrichment followed by deep sequencing (1.52 TB) was performed on 432 faecal samples from 181 IBD patients (CD=126;UC=55) starting biological therapy. Redundancy analysis and Dirichlet Multinomial Mixtures determined covariates of the virome composition and condensed the gut virota into viral community types.

**Results:** IBD patients were stratified based on unsupervised machine learning into two viral community types. Community type CA showed a low α-diversity and a high relative abundance of *<u>Ca</u>udoviricetes* [non-CrAss] phages and was associated to the dysbiotic Bact2-enterotype. Community type CrM showed a high α-diversity and a high relative abundance of *Caudoviricetes* [<u>Cr</u>Ass] and *<u>M</u>algrandaviricetes* phages. Gut virome variation was explained by patients’ individuality (75.8%), disease location (1.4%), age (0.5%) and faecal moisture (0.3%), with diagnosis not showing a non-redundant effect. Endoscopic outcome (0.5%) was accompanied by gut virome shifts in UC. Non-remitting UC, but not CD, patients revealed a high percentage of community type CA, a low diversity, and a high lysogenic potential. During pre-interventional analysis, we discovered five novel phages with a predictive value for therapeutic outcome.

**Conclusion:** The gut virota shows the existence of distinct virome configurations that are associated with endoscopic outcome, and community typing could be a valuable tool to improve our understanding about IBD subtypes, pathology, and activity.

**Significance of this study:** **What is already known on this subject?**

- Bacterial community typing (‘enterotyping’) is an established practice to stratify individuals based on their bacteriome composition
- IBD pathology is repeatedly associated with alterations in the gut virome reflected by a high *Caudovirales* and low *Microviridae* abundance
- IBD pathology is associated with a shift in phage lifestyle towards a highly lysogenic state of the gut virome

**What are the new findings?**

- Viral community types exist and could stratify individuals based on their gut virome composition in a large prospective multi-therapeutic IBD cohort
- Viral community types could be associated to the dysbiotic Bact2-enterotype
- Endoscopic outcome (remission/non-remission) was a significant covariate of the gut virome composition post-intervention and could be associated with viral community types in UC patients
- Non-remitting UC patients revealed a low diversity and a high abundance of lysogenic phages
- We discovered five novel phages (e.g., novel CrAss-like phage) associated with predicting treatment success

**How might it impact the clinical practice in the foreseeable future?**

- Viral community types could be a valuable tool to investigate IBD pathology and to differentiate between IBD subtypes and disease activity
- Viral community typing might distinguish a healthy gut from a dysbiotic gut virome
- Clinical studies should validate phages as predictive biomarkers in IBD patients starting therapy

## INTRODUCTION

The gut microbiota is a complex ecosystem that consists of viruses, fungi, bacteria, archaea, and protozoa. It can exert beneficial functions to the human host, such as protection against invading pathogens or production of essential vitamins. At times, this microbial ecosystem gets disrupted, resulting in gut dysbiosis. Gut dysbiosis is associated with several diseases, one of which is inflammatory bowel disease (IBD)^1^. IBD is a group of chronic remitting diseases involving inflammation of the gut, and its two main phenotypes include ulcerative colitis (UC) and Crohn’s disease (CD). Although the aetiology of IBD is unknown, the interplay between host genetic susceptibility, a mucosal immune response to the host microbiota and other environmental factors, has been suggested as working hypothesis^2^.

In recent years, the bacterial component of the microbiota has been repeatedly associated with the pathology and activity of IBD^3–5^. The gut microbiota of active IBD patients is characterized by a high abundance of *Proteobacteria* and a low abundance of *Firmicutes*, combined with a low bacterial α-diversity and cell count^2, 6^. Another frequently reported alteration is the reduction in anti-inflammatory bacteria, particularly butyrate-producing bacteria (e.g. *Faecalibacterium prausnitzii*)^7^. Community analysis can provide a more holistic view of the gut microbiota by collapsing the microbial variation into just a few categories. Enterotyping (or bacterial community typing) is such an analysis that can stratify patients based on their gut microbiota^8^. Four enterotypes have been reported, named *Bacteroides*1 (Bact1), *Bacteroides*2 (Bact2), *Prevotella* (Prev) and *Ruminococcus* (Rum)^6^. One enterotype, Bact2, largely reflects the IBD-specific bacterial alterations, as describe above^9^. Up to 80% of IBD patients harbor the Bact2-enterotype, which is seen as a dysbiotic enterotype, while less than 15% of healthy individuals possess this enterotype^6, 10^.

Despite the vast number of associations made with bacteria, little is known about the role of the viral component in the pathology and activity of IBD. A growing body of evidence suggests that disease pathology is associated with alterations in the gut virota as well^11–14^. These alterations are largely characterized by a high abundance of members of the *Caudovirales* and a low abundance of *Microviridae* members^15, 16^. Other alterations show a highly lysogenic potential of the gut virota ^11^. In addition, diversity changes have also been reported, albeit in an inconsistent manner^11–14, 17, 18^. One of the interesting viral groups is the recently identified CrAss-like phages, which are a diverse group that are believed to be the most abundant viruses present in the human gut^19^. Recently, Gulyaeva and colleagues described that CrAss-like phages are depleted in the IBD faecal virota^20^. Other studies have been suggesting that some lytic phages (e.g. CrAss-like phage crAss001) could exist in symbiosis with their bacterial host, and even drive bacterial diversity through a process called phase variation^21^. More specifically, phase variation allows the parallel multiplication of phages and their host by providing a balance between phage resistance and sensitivity. To date, no consistent associations have been made between the gut virota and disease activity. However, we anticipate that unravelling the full complexity of the human gut virome will deepen our understanding of complex human disease. Consequently, a viral counterpart of enterotyping (‘viral community-typing’) might improve this understanding and would allow stratification of individuals based on their gut virota.

In this study, we analyzed faecal samples (*n=432*) of a prospective cohort of active IBD patients (*n=181*) starting biological therapies and investigated the factors shaping the gut virome composition. Viral community typing was performed to describe the virome configurations, and to associate with covariates of the virome composition. In doing so, we are able to identify therapy outcome (as measured by endoscopic outcome) as a covariate of the gut virome, thereby highlighting the role of the gut virome in IBD.

## RESULTS

### The gut virome is dominated by *Caudoviricetes* and *Malgrandaviricetes* phages in a multi-therapeutic IBD cohort

Faecal samples were collected from patients in a prospective multi-therapeutic IBD cohort (*n=181*)^22^. Patients had either active ulcerative colitis (UC, *n=55*), or active Crohn’s disease (CD, *n=126*) and started biologicals as part of their medical care. Patients were re-evaluated at the pre-defined primary endpoint (post-intervention) and classified as achieving remission or not (Extended Data Figure 1; Extended Data Methods; Supplementary Table 1). The gut virome was characterized using the NetoVIR protocol to isolate, enrich and sequence viruses in faecal samples (Extended Data Figure 2a). Computational analyses on an input of 10.2 billion paired end reads (1.52 TB, *x̅*=23.6 million reads per sample) was performed with the latest methodologies (Extended Data Figure 2b). Most of the quality-controlled reads were found to be of viral origin (viral=63.0%, bacterial=32.3%, other=3.0%, dark matter=1.0%; Extended Data Figure 3). Most of the identified viruses could reliably be classified at class-level taxonomy (classified=93%, unclassified=7%; Extended Data Figure 4; Supplementary Table 2). The two most abundant viral classes were *Caudoviricetes* (58.1%, dsDNA tailed phages) and *Malgrandaviricetes* (37.7%, ssDNA circular phages), representing most quality-controlled reads (*x̅*=95.8%, range=2.76%-100% per sample). The former viral class could be broken down into *Caudoviricetes* [non-CrAss] and *Caudoviricetes* [CrAss] phages, encompassing 41.9% and 16.2% of the quality-controlled reads, respectively (Supplementary Table 2).

### The eukaryotic virome is small and is largely composed of plant viruses

A small percentage of samples contained eukaryotic viruses (32.9%) representing a minority of the quality-controlled viral reads (eukaryotic viruses=0.2%, phages=99.8%; Extended Data Figure 5; Supplementary Table 2). Eukaryotic viruses could be grouped based on the host, which could be known (animal, plant or fungal viruses) or unknown (small circular viruses). Most of the viruses detected in IBD patients belonged to the plant and fungal viral group, with only a few samples containing small circular viruses, or viruses potentially causing gastroenteritis. The two most prevalent viral species were Pepper mild mottle virus (prevalence=*12.9%,* genus=*Tobamovirus*) and Pepino mosaic virus (prevalence=*10.8%,* genus=*Potexvirus*), likely obtained via the patient’s diet (Supplementary Table 3)^23^.

### The gut virota reveals the existence of two virome configurations in IBD patients

The gut microbiota is complex and variable, further complicating its thorough exploration. One approach is to condense the bacterial complexity into bacterial community types (‘enterotypes’). By using Dirichlet Multinomial Mixture (DMM) modelling, bacterial research has consistently stratified large human gut microbiota studies into four enterotypes (Extended Data Methods). We used the same methodology and applied this for the first time to the gut virota. The gut virota observed in the IBD cohort consisted of no less than 874 genus-like groups (median=26, range=3-74 per individual). Applying the DMM algorithm reduced the viral complexity and revealed the existence of two distinct clusters, or virome configurations (*n=363*, genus-like group, Bray-Curtis dissimilarity; Figure 1a; Supplementary Table 4). The groups showed a high probability of cluster assignment and were hereafter referred to as viral community types (median=99.6%, Extended Data Figure 6; Supplementary Table 4).

**Figure 1.**
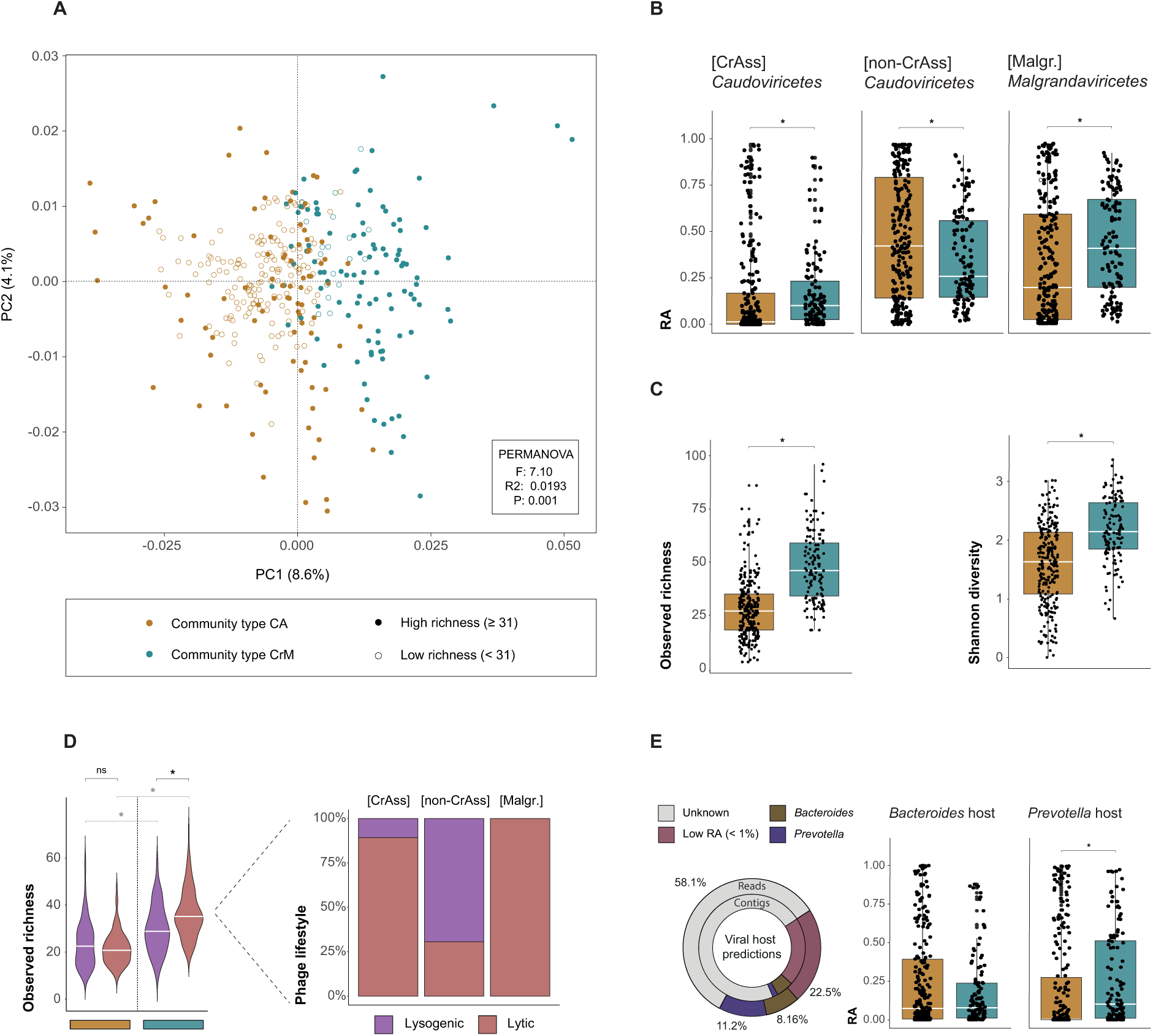
Viral community types to stratify individuals based on their virome composition. **A**, Principal coordinates analysis (PCoA) of inter-individual differences of the gut virome composition (genus-like group, Bray-Curtis dissimilarity) of the IBD cohort (coloured by viral community types, *n=363)* with shape representing categorical richness (median richness=31, open circles<31 (low richness), closed circles≥31 (high richness), R_2_=0.0193, P=0.001, Supplementary Table 8). **B**, Boxplot showing relative abundance of major phage classes (≥1% of reads) stratified according to viral community type (*n=363*, Mann-Whitney U, AdjP<0.05). **C**, Boxplot showing alpha-diversity metrics (Observed richness and Shannon diversity) stratified according to viral community type (*n=363*, Mann-Whitney U, AdjP<0.05). **D (left)**, Violin plot showing comparisons of observed richness stratified according to phage lifestyle (lytic versus lysogenic) within each viral community type (comparison within viral community type CrM, *n=121*, Mann-Whitney U, AdjP<0.05) and between viral community types (*n=363*, Mann-Whitney U, AdjP<0.05). **D (right)**, Barplot showing the distribution of phage lifestyles (lytic versus lysogenic) for major phage classes (≥1% of reads). **E (left)**, Donut plot visualizing host prediction of phages on contig and read level (≥1% of reads). **E (right)**, Boxplot visualizing host prediction of phages on read level (relative abundance) stratified according to viral community type (comparison *Prevotella*-infecting phages, *n=363*, Mann-Whitney U, AdjP<0.05). Adjustment for multiple testing (AdjP) was performed using the Benjamini-Hochberg method. Significant associations (AdjP<0.05) were visualized by an astrix (*). Abbreviations: Relative abundance (RA), Community type *Caudoviricetes* [non-CrAss] (CA) and community type CrAss-*Malgrandaviricetes* (CrM).

To obtain insights in these viral community types, we investigated the virome compositional variation and found that viral community types were associated with distinct groups of viruses (Figure 1b). The first viral community type (termed “CA”) revealed a high relative abundance of members of the *Caudoviricetes* [non-CrAss] (*n=363*, Mann-Whitney U, r=0.137, AdjP=0.0266; Supplementary Table 5), whereas the second (termed “CrM”) revealed a high relative abundance of members of the *Caudoviricetes* [CrAss] and *Malgrandaviricetes* (*n=363*, Mann-Whitney U, AdjP<0.05). Viral community types were characterized by a distinct α-diversity (Figure 1c). Viral community type CrM revealed a higher viral richness (*n=363*, Mann-Whitney U, r=0.542, AdjP<2.2e-16) and Shannon diversity (*n=363*, Mann-Whitney U, r=0.384, AdjP<4.76e-10) compared to viral community type CA. We hypothesized that this diversity variation might be the result of a different phage lifestyle (lytic versus lysogenic), as determined by the presence or absence of lysogeny-specific proteins such as integrases (Supplementary Table 6)^24^. We found that viral community type CrM, but not CA, was associated with a richness expansion of lytic compared to lysogenic phages (*n=121*, Mann-Whitney U, r=0.308, AdjP=6.86e-06; Figure 1d). In addition, only phages belonging to the *Caudoviricetes* class were capable of lysogeny (*Caudoviricetes* [non-CrAss]=69.4%, *Caudoviricetes* [CrAss]= 10.8%, *Malgrandaviricetes*=0%; Figure 1d). Overall, both community types showed a high relative abundance of lytic compared to lysogenic phages (Mann-Whitney U, AdjP<0.05; Supplementary Table 5), but only in viral community CrM we observed a high relative abundance of lytic *Caudoviricetes* phages (CrAss+non-CrAss, *n=121*, Mann-Whitney U, r=0.307, AdjP=0.00360; Supplementary Table 5), suggesting that *Caudoviricetes* prophages were induced in patients with viral community type CA.

Furthermore, viral community types were associated with IBD subtype (proportion test, X^2^=5.20 AdjP=0.0472; Supplementary Table 5) but not disease location (proportion test, X^2^=1.04 AdjP=0.791; Supplementary Table 5), as characterized by a higher prevalence of viral community type CA in CD compared to UC patients (CD=69.8%, UC=57.9%, Supplementary Table 5).

### Viral community type CA is mostly associated with the dysbiotic Bact2-enterotype

Next, we implemented *in silico* phage host prediction to determine the bacterial host. Most bacterial hosts were predicted on phylum-level (83.6%) and only a few hosts could be predicted on genus-level (27.7%; Extended Data Figure 7). The majority of the host phyla were *Bacteroidetes*, *Firmicutes* and *Proteobacteria*. Most of the host genera were *Bacteroides* and *Prevotella*, which were preferentially infected by *Caudoviricetes* [non-CrAss] and *Caudoviricetes* [CrAss] phages, respectively (≥1% reads; Figure 1e). Viral community type CrM revealed a high relative abundance of *Prevotella*-infecting phages compared to viral community type CA (*n=363*, Mann-Whitney U, r=0.235, AdjP=1.51e-05; Supplementary Table 5), but no significant differences were found in relative abundance of *Bacteroides*-infecting phages between community types (*n=363*, Mann-Whitney U, r=0.00530, AdjP=1.00; Supplementary Table 5).

The gut microbiota of IBD patients has been repeatedly associated with the dysbiotic Bact2-enterotype^9, 22^. Extended Data Figure 8 shows a statistical correlation between the bacterial enterotypes (Bact1, Bact2, Prev and Rum) and the viral community types. Viral community type CA was mostly associated with the Bact2-enterotype. This dysbiotic enterotype showed a higher prevalence in viral community type CA compared to viral community type CrM in UC (CA=57.1%, CrM=25.6%, *n=88*, proportion test, X^2^=7.55, AdjP=0.012; Supplementary Table 7) and in CD patients (CA=79.6%, CrM=38.3%, *n=207*, proportion test, X^2^=3.13, AdjP=8.96e-08). Conversely, viral community type CrM was mostly associated with the Bact1-enterotype. This enterotype showed a higher prevalence in viral community CrM compared to viral community type CA in UC (CA=16.3%, CrM=48.7%, *n=88*, proportion test, X^2^=9.24, AdjP=9.44e-03) and CD patients (CA=12.2%, CrM=35.0%, *n=88*, proportion test, X^2^=1.30, AdjP=4.20e-04).

### The gut virome composition is individual and is associated with disease location, patient’s age, and moisture content of the faecal samples

The factors shaping the gut virome composition were determined by distance-based redundancy analysis (dbRDA) in the IBD cohort (Supplementary Table 9). Patient individuality was identified as the largest explanatory variable, thereby reinforcing the notion that the virome is highly individual-specific (*n=363*, multivariate dbRDA, genus-like group, R_2_=75.8%, AdjP=0.001; Figure 2a). Disease location was identified as the second largest explanatory variable of virome variation (*n=363*, multivariate dbRDA, genus-like group, R_2_=1.40%, AdjP=0.001). Next, patient age and moisture content of the stool also showed a limited contribution to the virome variation (*n=363*, multivariate dbRDA, genus-like group, age R_2_=0.5%, moisture R_2_=0.3%, AdjP<0.05). Remarkably, disease location (Montreal classification) stratifying patients according to ileal (L1CD), colonic (L2CD/UC) and ileocolonic (L3CD) phenotypes, revealed a higher explanatory power than simple diagnosis (UC/CD), and was observed in bacterial research as well (*n=363*, univariate dbRDA, genus-like group, location R_2_=1.34%, diagnosis R_2_=0.47%, AdjP<0.05)^22^.

**Figure 2.**
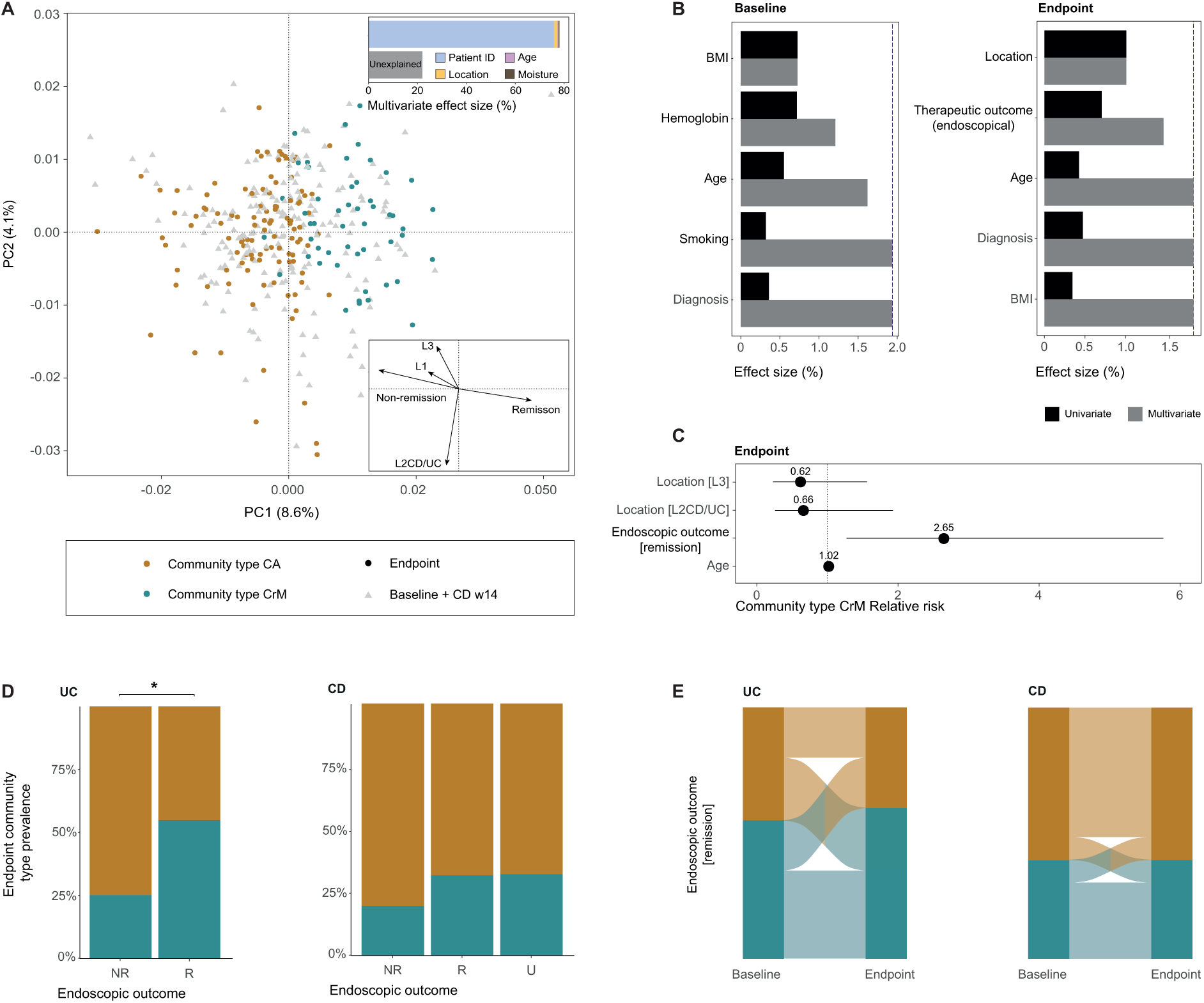
Virome covariates of the IBD cohort, and their association to viral community types. **A**, Principal coordinate analysis of inter-individual differences of the gut virome composition (genus-like group, Bray-Curtis dissimilarity) of the IBD cohort (coloured by viral community types, *n=363*) with shape representing sample timepoint. Insert top: Barplot represents significant covariates of virome composition of the IBD cohort, as identified in a multivariate model. The cumulative model (patient ID, age, location and moisture content) explains 78.0% of the virome variation (stepwise multivariate R_2_). Arrows represent the effect sizes of a post-hoc fit of significant virome covariates of post-intervention samples, as identified in a multivariate model in B (right). **B**, Metadata variables correlating (driving) significantly to virome variation in (left) pre-intervention and (right) post-intervention samples (dbRDA, genus-like group, Bray-Curtis dissimilarity). Effect sizes of correlating metadata are calculated independently (univariate coloured in black, redundant covariates) or in a cumulative model (multivariate coloured in black, non-redundant covariates). The purple dashed line represents the threshold for significant contribution to the multivariate model (metadata variables exceeding threshold coloured in black). **C, D, E,** Modeling the association between the prevalence of viral community type CrM and significant non-redundant covariates (location, therapeutic outcome as measured by endoscopy and age) of post-intervention samples (logistic regression model, *n=166*). **C**, Relative risk ratio of IBD patients hosting community type CrM associated with significant non-redundant covariates of the virome composition in post-intervention samples (endoscopic outcome [remission], *n=166*, RR=2.65, AdjP<0.05). **D**, Barplot showing viral community type (CA/CrM) prevalence in post-intervention samples stratified according to endoscopic outcome (R, NR, U) for UC (left) and CD (right) patients (comparison UC endoscopic outcome, *n=51*, proportion test, AdjP<0.05). **E**, Alluvial diagram showing transitions of viral community types after successful intervention for UC (left) and CD (right) patients (logistic regression, *n=79×2,* AdjP>0.05). A total of 25.0% of UC patients and 9.09% of CD patients shift towards viral community type CrM, whilst only 20.0% of UC patients and 9.09% of CD patients shift towards viral community type CA. Adjustment for multiple testing (AdjP) was performed using the Benjamini-Hochberg method. Significant associations (AdjP<0.05) were visualized by an astrix (*). Abbreviations: Ulcerative colitis (UC), Crohn’s disease (CD), Remission (R), Non-remission (NR), Unknown (U), pre-intervention (baseline), post-intervention (Endpoint/primary endpoint), Community type *Caudoviricetes* [non-CrAss] (CA) and community type CrAss-*Malgrandaviricetes* (CrM).

To further evaluate the factors shaping the gut virome composition in active IBD patients, we focused on baseline (pre-intervention) samples (Figure 2b; Supplementary Table 10). We identified body-mass index (BMI), haemoglobin concentration, age, and smoking behaviour as unique contributors to gut virome variation in baseline samples (*n=151*, multivariate dbRDA, genus-like group, R_2_=1.94%, AdjP=0.050, full model). We found that the patient’s BMI correlated with hemoglobin concentration (*n=151*, ρ=0.230, AdjP=0.00460; Supplementary Table 11) and reasoned that a low BMI and a low hemoglobin concentration (anaemia) probably expressed frailty of active IBD patients. In addition, we hypothesized that the gut virome might have a predictive capacity to determine therapeutic outcome (remission or non-remission at primary endpoint). Unfortunately, none of the disease activity indices could be associated to virome composition at baseline; however, viral community types could be associated to endoscopic outcome (*n=151*, AdjP<0.050; Supplementary Table 10). Here, remitting patients harboured a 225% increased probability of hosting viral community type CrM, while simultaneously presenting a poor predictive power (univariate logistic regression, AUC=60.0, AdjP=0.0348; Supplementary Table 10), suggesting that community types had slight pharmacodynamic capabilities able to track patient outcome.

### The gut virome composition and community types are associated with endoscopic outcome in UC patients

To determine the effect of therapy outcome on the gut virome composition we investigated the association of all covariates on primary endpoint (post-intervention) samples in IBD patients (Figure 2b; Supplementary Table 12). Disease location and patient’s age were once more identified as unique explanatory variable (*n=166*, univariate dbRDA, genus-like group, Location R_2_=1.01%, Age R_2_=0.370 %, AdjP<0.050). Interestingly, therapeutic outcome appeared as a new covariate of the gut virome composition, as measured by the golden standard of endoscopic evaluation (*n=166*, multivariate dbRDA, genus-like group, R_2_=0.460%, AdjP=0.0320). On the other hand, other disease activity indices (clinical and biomarker evaluation) did not have a significant contribution to the virome variation. Moreover, no association was found between remission rates and disease location (*n=138*, proportion test, X^2^=7.47, r=0.233, AdjP=0.0584; Supplementary Table 13), most likely because of limited samples sizes.

By focusing on post-intervention samples, this study premiered endoscopic outcome as a significant covariate of the gut virome composition in IBD patients. We hypothesized that virome covariates might be associated to viral community types in post-intervention samples, and therefore modelled the association between viral community types and the three significant covariates of virome variation (*n=166*, logistic regression; Figure 2c; Supplementary Table 14). The disease location and patient’s age could not be associated to viral community types (*n=166*, AdjP>0.05), while the endoscopic outcome could be associated to viral community types (*n=166*, AdjP=0.0280). Remitting patients harboured a 265% increased probability of hosting viral community type CrM (endoscopic remission relative risk (RR)=2.65) and a 62% decreased probability of hosting viral community type CA (endoscopic non-remission RR=0.38). Stratification of endoscopic outcome for each IBD subtype demonstrated that UC, but not CD patients, had a higher prevalence of viral community type CrM in remitting patients (Figure 2d). Viral community type CrM was found in 54.8% of the remitting and 25.0% of the non-remitting UC patients (*n=51*, proportion test, X^2^=4.41, r=0.254, AdjP=0.0357; Supplementary Table 15). Conversely, viral community type CrM was found in 31.7% of the remitting and 19.6% of the non-remitting CD patients (*n=115*, proportion test, X^2^=2.21, r=0.136, AdjP=0.345; Supplementary Table 15). Next, we evaluated the impact of treatment success (endoscopic outcome) on the gut virome, as determined by compositional changes between baseline and primary endpoint (paired statistics, *n=2×103*, Supplementary Table 9). There were no changes in the gut virome composition over time associated with endoscopic remission, across all patients (paired dbRDA, n=2×53, R_2_=0.869%, AdjP=0.688), or following the stratification of the patients into corresponding UC and CD subtypes (Figure 2e; Supplementary Table 14).

### Intestinal inflammation in UC patients is associated with a low viral diversity and a high lysogenic potential of *Caudoviricetes*

To investigate the effect of the endoscopic outcome on the major viral classes in more detail we focused on post-interventional samples of the IBD patients. The major viral classes of UC, but not CD patients, were characterized by a distinct α-diversity between remitting and non-remitting patients (Figure 3a,c; Supplementary Table 16-18). Non-remitting UC patients revealed a low Shannon diversity of *Caudoviricetes* [non-CrAss] (*n=51*, Mann-Whitney U, r=0.372, AdjP=0.0143; Supplementary Table 16) and *Malgrandaviricetes* (*n=51*, Mann-Whitney U, r=0.329, AdjP=0.0378; Supplementary Table 16) phages compared to remitting UC patients. Other metrics (Pielou’s evenness and richness) were not seen to be affected by the endoscopic outcome. Conversely, none of the metrics (Shannon diversity, Pielou’s evenness and richness) were affected by the endoscopic outcome in CD patients (*n=94*, Mann-Whitney U, AdjP<0.05; Supplementary Table 16-18). Next, we hypothesized that therapeutic outcome might be associated to a changing phage lifestyle (Figure 3b,c). A high relative abundance of lysogenic phages was observed in non-remitting compared to remitting UC patients (*n=51*, Mann-Whitney U, r=0.335, AdjP=0.0344), while no differences in phage lifestyle were found between non-remitting and remitting CD patients (*n=94*, Mann-Whitney U, r=0.219, AdjP=0.0677; Supplementary Table 19). Moreover, the relative abundance of lysogenic phages in (non)-remitting UC patients was not associated to viral community types (*n=20*, Mann-Whitney U, r=0.127, AdjP=1.00; Supplementary Table 19). Therefore, the expanded lysogenic potential of phages was thought to be associated with endoscopic outcome rather than viral community types. The high lysogenic potential in non-remitting UC patients appeared to be associated with the entire class of *Caudoviricetes* phages ([CrAss + non-CrAss], *n=51*, Mann-Whitney U, r=0.313, AdjP=0.0498; Supplementary Table 20). To evaluate the finding by Gulyaeva and colleagues that CrAss-like phages in the human gut were depleted in IBD patients, we also assessed the association between the prevalence of *Caudoviricetes* [CrAss] and endoscopic outcome^20^. UC patients showed a prevalence of 77.4% in remission compared to 60.0% in non-remission (*n=51*, proportion test, X^2^=1.04, AdjP=0.617; Supplementary Table 21). CD patients showed a prevalence of 71.9% in remission compared to 62.1% in non-remission (*n=94*, proportion test, X^2^=1.76, AdjP=0.419). In addition, no associations were found between the relative abundance of *Caudoviricetes* [CrAss] and endoscopic outcome.

**Figure 3.**
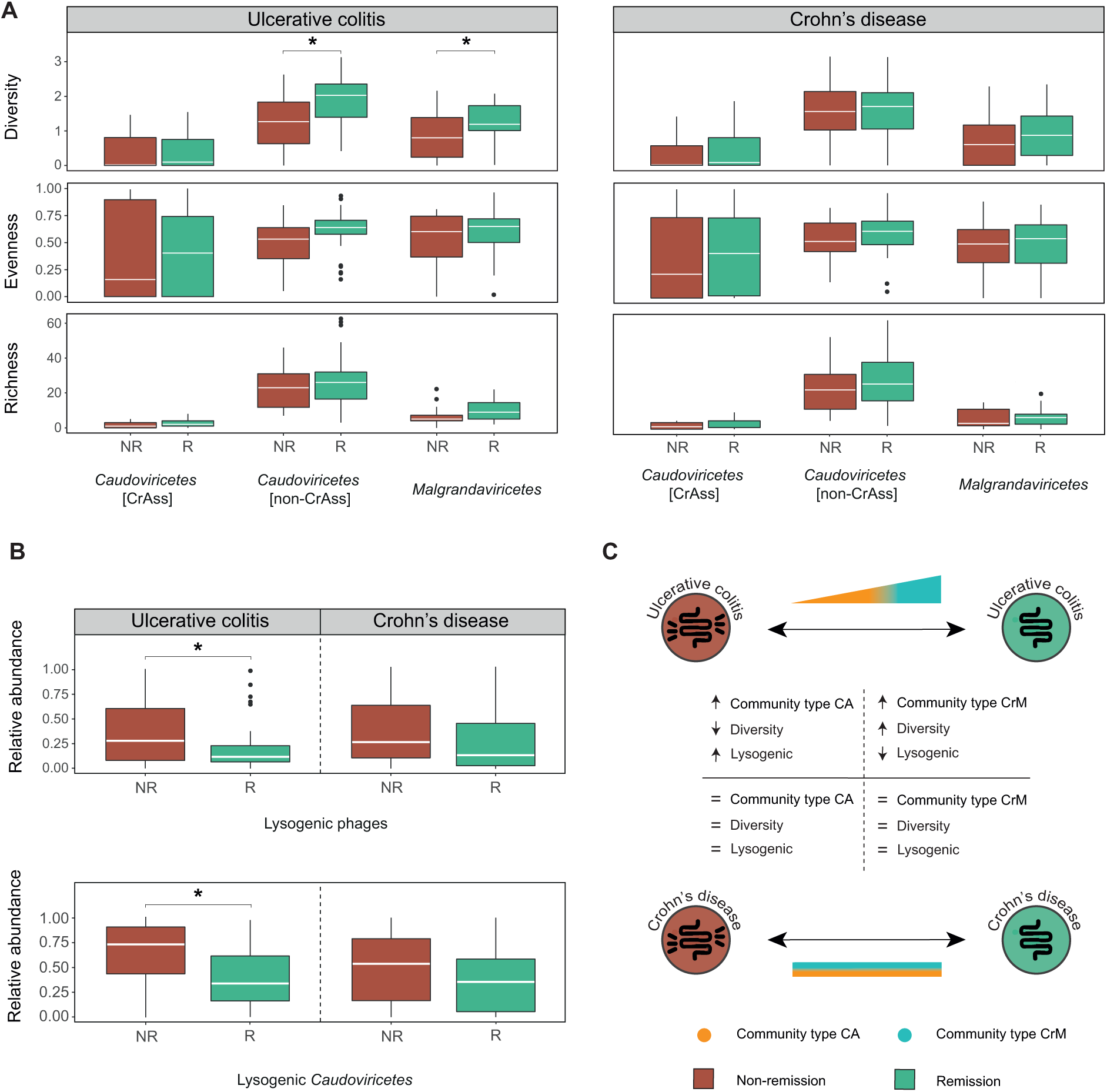
Virome characteristics of post-intervention samples stratified according to endoscopic outcome. **A**, Boxplot showing alpha-diversity metrics (Observed richness and Shannon diversity) and evenness (Pielou’s evenness) for major phages classes (> 1% reads) stratified according to endoscopic outcome (NR/R) for UC (left) and CD (right) patients (comparison *Malgrandaviricetes* and *Caudoviricetes* [non-CrAss] diversity in UC, Mann-Whitney U test, AdjP<0.05). **B**, Boxplot showing relative abundance of lysogenic (top) and lysogenic *Caudoviricetes* phages stratified according to endoscopic outcome (NR/R) for UC (left) and CD (right) patients (comparison lysogenic (*Caudoviricetes* [CrAss + non-CrAss]) phages in UC, Mann-Whitney U test, AdjP<0.05). **C**, Graphical summary of virome characteristics detected in post-intervention samples. Adjustment for multiple testing (AdjP) was performed using the Benjamini-Hochberg method. Significant associations (AdjP<0.05) were visualized by an astrix (*). Abbreviations: Remission (R), Non-remission (NR), Community type Caudoviricetes [non-CrAss] (CA) and community type CrAss-Malgrandaviricetes (CrM).

### Phages have a predictive capacity to determine therapeutic success

To investigate the predictive capacity of individual phages in determining therapeutic success, we focused on pre-interventional samples of IBD patients. We argue that phages with an appropriate predictive capacity should be (1) highly shared between patients and (2) differentiate between endoscopic treatment success (remission versus non-remission). Firstly, the 20 most prevalent viruses were determined for each IBD subtype and at least one of them was found in 97.7% of UC and 87.6% of CD patients (Extended Data Figure 9, Supplementary Table 22). Secondly, phages with a capability to differentiate between endoscopic treatment success were identified for each IBD subtype (LDA score (log10) > 2, AdjP<0.05; Figure 4a, Supplementary Table 23). Accordingly, five individual novel phages (both highly prevalent and differentially present) were found with potential predictive capacity (*n_UC_ =44*, *n_CD_*=99; Figure 4b; Supplementary Table 24-25). Specifically, UC patients harboured two phages predicting endoscopic remission (CrAssella-R and Cripes-R) and one phage predicting endoscopic non-remission (Croccus-NR). On the Contrary, CD patients harboured two phages predicting endoscopic remission (CrAssella-R and Croides-R) and one phage predicting endoscopic non-remission (Croides-NR). CrAssella-R is a *Caudoviricetes* [CrAss] phage that was associated with treatment success and the only phages detected in both of the IBD subtypes. We argued that next-generation sequencing methods, in combination with stringent bioinformatic criteria might underestimate the presence and quantity of phages in IBD patients. Therefore, qPCR was used to quantify and validate the potential predictive capacity of the five above mentioned phages (Supplementary Table 26). Consequently, two phages in UC with potential predictive capacity (Cripes-R and Croccus-NR) revealed a higher qPCR positivity rate than NGS positivity rate (*n=44*, qPCR=79.5%/79.5%, NGS=16.9%/18.2%, respectively; Figure 4c; Supplementary Table 27). Three phages in CD with potential predictive capacity (CrAssella-R, Croides-R and Croides-NR) revealed a higher qPCR positivity rate as well (*n=88*, qPCR=21.4%/52.4%/36.4%, NGS=16.2%/21.2%/10.1%; Supplementary Table 27). Only one phage with a potential predictive capacity (CrAssella-R) revealed a slightly lower qPCR positivity rate (*n=44,* qPCR=22.7%, NGS=29.5%). Next, the true predictive capacity (IBD predictive value [IPV]) of individual phages was determined based on the viral copy number (Supplementary Table 27). Remitting IBD patients were predicted by a positive IPV (*n=82*, IPV>0, sensitivity=0.67; Figure 4d; Supplementary Table 27) and non-remitting IBD patients were predicted by a negative IPV (*n=82*, IPV<0, specificity=0.68). No predictions were made for IBD patients with a neutral IPV (*n=46,* IPV=0). Next, we modelled the association between prevalence of remission and IPV and found a predictive power of 72.8% (*n=82, l*ogistic regression, AUC=72.8%, AdjP=0.000394; Figure 4e; Supplementary Table 28). At last, we assessed a combination of IPV and viral community types and found that the latter variable does not significantly contribute to the model. In general, viruses revealed for the first time predictive capacity in determining therapeutic success in IBD patients, in which CrAss-like phages play an important role.

**Figure 4.**
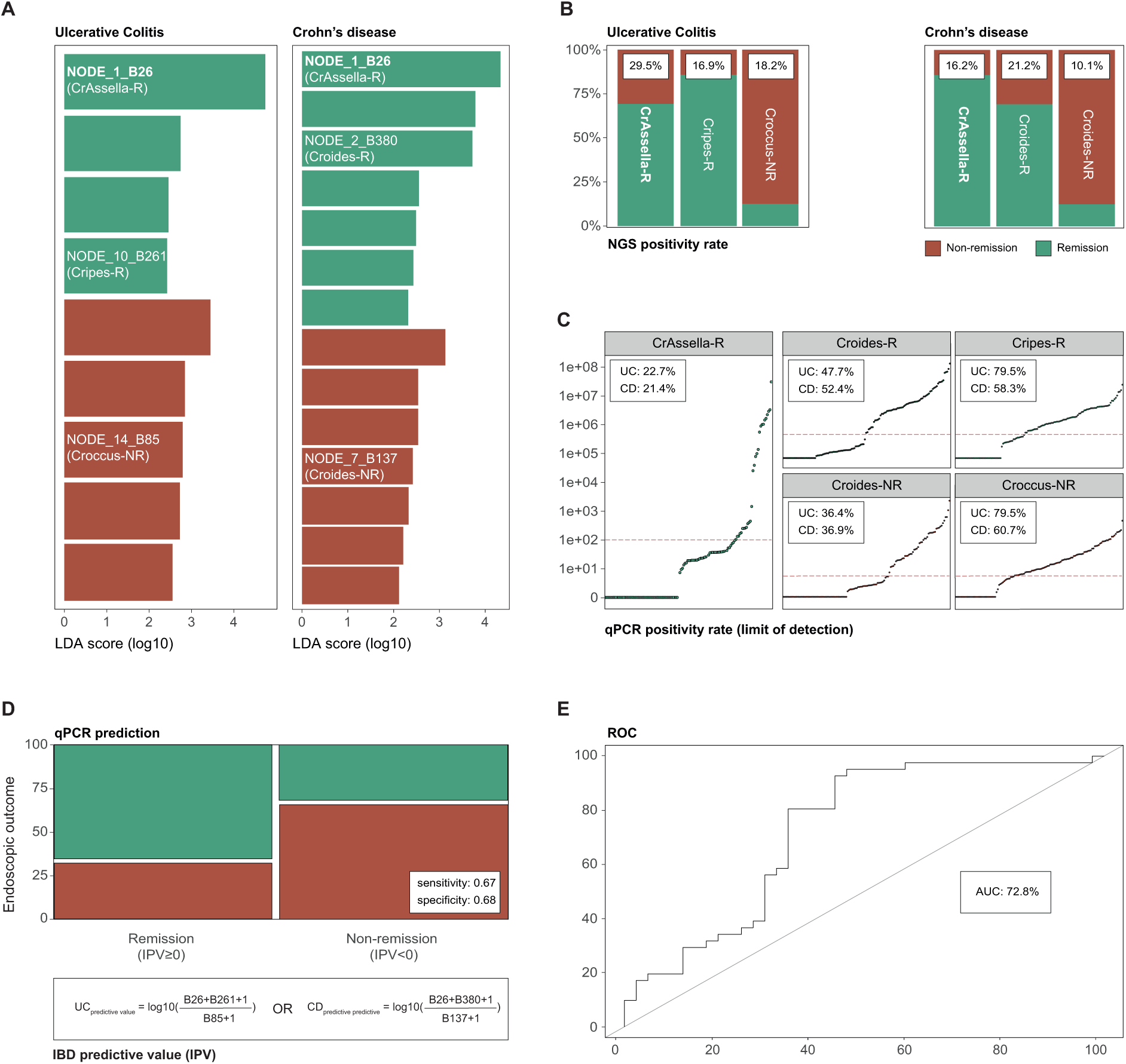
Predictive potential of the individual gut phages to determine therapeutic (endoscopic) outcome in IBD patients undergoing biological therapy. **A**, Pre-interventional (baseline) differential abundance analysis of individual gut viruses to determine endoscopic outcome (non-remission/remission) by linear discriminant analysis (LDA) and effect size (Lefse) in IBD patients undergoing biological therapy (LDA score (log10) ≥ 2, *n=143*, Mann-Whitney U test, AdjP<0.05). The NODE names of viruses with potentially predictive capacity for determining endoscopic outcome within UC (left, *n=44*) or CD (right, *n=99*) patients are coloured in white. **B**, Barplot representing the prevalence of five phages with potential predictive capacity as stratified for endoscopic outcome for UC (left, three phages) and CD (right, three phages) patients. NODE_1_B26 is found to have predictive capacity for both IBD subtypes (bold). NGS positivity rate (prevalence in baseline samples) is shown for each phage in the white boxes (*n=143*). **C**, Scatterplot representing the number viral copies in baseline samples for each potential predictive phage (*n=128*). qPCR positivity rate (prevalence in baseline samples) is shown for each predictive phage in the white boxes (limit of detection ≥ 100 viral copies, represented by red dashed line) for IBD subtypes. **D**, **E**, The predictive capacity of individual phages to determine endoscopic outcome is measured by the concept of IPV based on viral copy number. IPV is calculated by the natural logarithm of the remission predictive phages over the non-remission predictive phages for IBD subtypes (bottom formula, limit of quantification ≥ 500 viral copies). **D**, Mosaic plot displaying the distribution of (excluding samples with no predictions) the categorical variables endoscopic remission and qPCR prediction (based on IPV, sensitivity=0.67, specificity=0.68). **E,** Modelling the association between prevalence of remission and IPV (logistic regression, *n=88,* RR=1.32, AdjP<0.05). ROC curve representing predictive potential of phages (*n=88*, AUC=72.8%). Adjustment for multiple testing (AdjP) was performed using the Benjamini-Hochberg method. Abbreviations: IBD predictive value (IPV), Ulcerative colitis (UC), Crohn’s disease (CD), Inflammatory bowel disease (IBD), Next-generation sequencing (NGS) and Receiver operating characteristic (ROC) curve.

## DISCUSSION

To date, this research represents the most comprehensive gut virome study (using viral particle enrichment followed by deep sequencing) of IBD patients undergoing biological treatment. Here, we implemented the concept of viral community typing and found that patients could be stratified into two (CA/CrM) virome constellations (Figure 1a). We hypothesize that, patients harbouring viral community type CA exhibited a dysbalanced virome, characterised by a high abundance of *Caudoviricetes* [non-CrAss] and low viral diversity, which has been observed before (Figure 1b,c)^12^. The association with the dysbiotic Bact2-enterotype strengthens the belief that this constellation reflects the viral counterpart of gut dysbiosis (Extended Data Figure 8). Conversely, patients harbouring viral community CrM might exhibit a more eubiotic-like virome characterised by a high abundance of members of the *Caudoviricetes* [CrAss] and *Malgrandaviricetes*, a high viral diversity and a shift away from the dysbiotic Bact2-enterotype.

Furthermore, remitting UC patients are associated to viral community type CrM, whereas non-remitting UC patients are associated to viral community type CA, thereby reinforcing the notion that viral community types might reflect a potential dysbalanced or eubiotic-like state of the gut virome, and may therefore be clinically relevant (Figure 2b-d). However, these findings could not be reproduced for CD patients suggesting that these patients retain a dysbalanced state even in the case of remission or quiescent disease. Thus far, twin studies revealed a larger genetic impact in CD pathology and in contrast, a larger non-genetic or environmental (e.g., virome) impact in UC pathology^25^. These findings are corroborated in this study, further revealing a substantial role of the gut microbial ecosystem in UC pathology (Figure 3). Consequently, the thorough analysis of the gut inflammation in IBD revealed a decrease in viral diversity and an expansion of lysogenic phages. However, changes in viral diversity and phage lifestyle were not observed in CD pathology, thereby once more confirming the different role of the virome regarding the IBD subtype.

We argue that a persistent inflamed state of the intestine will shape the gut virome by acting on different mechanisms. Firstly, an increase in gut motility might lower the microbial (and viral) diversity due to an increase defecation frequency^26^. Secondly, phage-mediated lysis might explain the observed expansion of lysogenic gut phages^27^. Phage-mediated lysis describes a positive-feedback loop between phage induction and intestinal inflammation. Briefly, inflammation stimulates enterocytes to produce stressors (eg. reactive oxygen species) activating a stress response in the host bacteria (‘SOS response’). The stress response will trigger prophages to initiate the lytic lifecycle, leading to the lysis of the bacterial host cell. An increased bacterial lysis will be accompanied by an increase of pathogen-associated molecular patterns (e.g., lipopolysaccharide, bacterial DNA) thar are stimulating pattern recognition receptors on enterocytes. These cells will produce more stressors and further promote prophage induction, thus, starting a positive feedback-loop and increase the lysogenic potential under intestinal inflammation. Taken together, these mechanisms might explain the observed decrease in viral diversity and lysogenic expansion under inflammatory conditions (Figure 3).

Following the viral exploration of the gut inflammation, we also discovered five novel phages that were associated with predicting treatment success, as confirmed by qPCR results (Figure 4). One phage, a novel CrAss-like phage (CrAssella-R), revealed predictive capacity for both IBD subtypes. *Shkoporov* and colleagues described that long-term persistence of crAss0001 with its bacterial host could drive diversity (hallmark of eubiosis) by a process called phase variation^21^. We argue that biological treatment induces remission and that mechanism such as phase variation could provide a path to maintain remission by stimulating eubiosis. Phages with predictive abilities might be those capable of driving diversity in combination with the respective host.

In conclusion, in this study we have shown that, viral community typing allows the stratification of IBD patients based on a distinct viral composition in the gut, and could be used to better understand IBD subtypes, pathology and disease activity in the future.

## METHODS

Faecal samples were collected from a prospective IBD cohort (126 CD and 55 UC) based on sample availability and pairing (*n=432*). All IBD patients had active disease at baseline defined by endoscopy and were started on one of four approved biological therapies (infliximab, adalimumab, ustekinumab or vedolizumab). Every patient provided a baseline and a follow-up (primary endpoint) sample. The NetoVIR protocol was used to prepare faecal samples for viral metagenomics, as described before (Extended Data Figure 2A)^28^. Further bioinformatic processing, viral community typing, diversity analysis and other details regarding the protocols were elaborately described in the attachment (Extended Data Methods).

### Supplementary Tables (in excel file, with full legend on first tab)

**Supplementary Table 1.** Metadata descriptions of sample selection of prospective cohort of IBD samples undergoing biological therapy (*n=432*).

**Supplementary Table 2.** Characteristics of the rarefied gut virome in a prospective IBD cohort (*n=377*)

**Supplementary Table 3.** Characteristics of the most common eukaryotic viruses detected in a prospective IBD cohort (*n=377*)

**Supplementary Table 4.** Cluster (viral community type) assignment results from dirichlet multinomial (DMM) clustering (*n=363*)

**Supplementary Table 5.** Cluster (viral community type) CHRACTERISTICS. Alpha diversity (Richness and Shannon index), Relative abundance of phages classes, phage lifestyle, host prediction, location and diagnostic discrepancies between DMM clusters (n=363, Mann-Whitney U, adjusted for multiple testing (AdjP, BH method))

**Supplementary Table 6.** Genes marking lysogenic lifestyle as detected by Cenote-Taker2

**Supplementary Table 7.** Association between viral community types and enterotypes (*n*=295, proportion test, adjusted for multiple testing (AdjP, BH method))

**Supplementary Table 8.** Permutational MANOVA Adonis test of categorical richness (*n*=363, Bray-Curtis dissimilarity)

**Supplementary Table 9.** Covariates with non-redundant explanatory power on the genus level ordination of the complete IBD cohort, determined by univariate and multivariate distance-based redundancy analysis (single/paired) at genus level (*n=363*, dbRDA, Bray-Curtis dissimilarity, adjusted for multiple testing (AdjP, BH method))

**Supplementary Table 10.** Covariates with non-redundant explanatory power on the genus level ordination of pre-intervention samples, determined by univariate and multivariate distance-based redundancy analysis at genus level (*n=151*, dbRDA, Bray-Curtis dissimilarity, adjusted for multiple testing (AdjP, BH method)). Predictive model (logistic regression modelling) of binary response variable (viral community types) using endoscopic outcome in pre-intervention samples (n=128, glm with binomial link, adjusted for multiple testing (AdjP, BH method)).

**Supplementary Table 11.** Association between BMI and hemoglobin concentration of pre-intervention (*n=151*, Spearman correlation)

**Supplementary Table 12.** Covariates with non-redundant explanatory power on the genus level ordination of post-intervention samples, determined by univariate and multivariate distance-based redundancy analysis at genus level (*n=166*, dbRDA, Bray-Curtis dissimilarity, adjusted for multiple testing (AdjP, BH method))

**Supplementary Table 13.** Association between endoscopic outcome and disease location of post-intervention samples (n=138, proportion test, adjusted for multiple testing (AdjP, BH method))

**Supplementary Table 14.** Logistic regression modelling of binary response variables (community type CrM) in post-intervention samples using single non-redundant explanatory variables (*n=166*, glm with binomial link, adjusted for multiple testing (AdjP, BH method))

**Supplementary Table 15.** Associations between categorical variable (endoscopic outcome) and community type distribution with post-hoc pairwise comparisons for > 2 groups (proportion test, adjusted for multiple (AdjP, BH method))

**Supplementary Table 16.** Alpha diversity (Shannon index) comparisons of phage classes in post-intervention samples (Mann-Whitney U test, adjusted for multiple testing (AdjP, BH method))

**Supplementary Table 17.** Pielou’s evenness comparisons of phage classes in post-intervention samples (Mann-Whitney U test, adjusted for multiple testing (AdjP, BH method))

**Supplementary Table 18.** Alpha diversity (Observed richness) comparisons of phage classes in post-intervention samples (Mann-Whitney U test, adjusted for multiple testing (AdjP, BH method))

**Supplementary Table 19.** Association between relative abundance of phage lifestyle and endoscopic outcome in post-intervention samples (Mann-Whitney U test, adjusted for multiple testing (AdjP, BH method))

**Supplementary Table 20.** Association between relative abundance of phage lifestyle of *Caudoviricetes* phages and endoscopic outcome in post-intervention samples (Mann-Whitney U test, adjusted for multiple testing (AdjP, BH method))

**Supplementary Table 21.** Prevalence of CrAss-like phages in post-intervention samples within disease (proportion test, adjusted for multiple testing (AdjP, BH method))

**Supplementary Table 22.** Detailed description of the top 20 most prevalent phages for each disease subtype at baseline (*N=143*).

**Supplementary Table 23.** Differential abundant viruses at baseline between endoscopic outcome (remission/non-remission) were determined using linear discriminant analysis (LDA) effect size (Lefse) (*n=143*, Mann-Whitney U test, adjusted for multiple testing (AdjP, BH method))

**Supplementary Table 24.** NGS positivity rate of potential predictive biomarkers, and comparisons of ngs positivity rate of potential predictive features between endoscopic outcome(remission/non-remission) at baseline (*n=143*, proportion test, adjusted multiple testing (AdjP, BH method))

**Supplementary Table 25.** Detailed description of predictive phage markers at baseline

**Supplementary Table 26.** QPCR primers, probes, standards and conditions of potential predictive baseline phage markers

**Supplementary Table 27.** QPCR positivity rate of potential predictive biomarkers for each disease subtype (> 500 viral copies per mL, *n=128*).

**Supplementary Table 28.** Contingency table and logistic regression modelling of binary response variables (endoscopic outcome) in the baseline IBD cohort using IBD predictive value input variables (*n=82*, glm with binomial link, adjusted for multiple testing (AdjP, BH method))

## Data Availability

All data produced in the present work are contained in the manuscript

## Acknowledgements

We would like to thank all participating patients for their contribution. We thank Nooshin Ardeshir Davani for technical support regarding faecal sample logistics and Prof. Dr. Jurgen Vercauteren for the support in statistical analysis. We thank Prof. Dr. Evelien Adriaenssens for assisting in the nomenclature of the phages with predictive potential and the VIB nucleomics Core for sequencing production (www.nucleomics.be). We thank Winston Chiu for extensive proofreading of the manuscript. This research was supported by the ‘Fonds Wetenschappelijk Onderzoek’ (Research foundation Flanders) (Daan Jansen: 1S78021N; Kathleen Machiels: 12M9118N). Kathleen Machiels was part of this work as a postdoctoral fellow and is employed as medical advisor in Pfizer since 05/01/2021.

## Author Contributions

The study was conceived by JM, SV and JR. Experiments were designed by DJ, SV, JR and JM. Sampling was set up by CC, KM and SV. Experiments were performed by DJ, TM and CS. Bioinformatic and statistical analysis of the sequences reads was performed by DJ, SS and GF. DJ, CS, GF, SS and JM drafted the manuscript. All authors revised the article and approved the final version for publication.

## Author information

SV has received grants from AbbVie, J&J, Pfizer, Galapagos, Takeda. SV has received consulting and/or speaking fees from AbbVie, AbolerIS Pharma, AgomAb, Alimentiv, Arena Pharmaceuticals, AstraZeneca, Avaxia, BMS, Boehringer Ingelheim, Celgene, CVasThera, Dr Falk Pharma, Ferring, Galapagos, Genentech-Roche, Gilead, GSK, Hospira, Imidomics, Janssen, J&J, Lilly, Materia Prima, MiroBio, Morphic, MrMHealth, Mundipharma, MSD, Pfizer, Prodigest, Progenity, Prometheus, Robarts Clinical Trials, Second Genome, Shire, Surrozen, Takeda, Theravance, Tillots Pharma AG, Zealand Pharma. Other authors report no conflict of interest. Readers are welcome to comment on the online version of the paper. Correspondence should be addressed to JM.

## Patients and Public involvement statement

The Leuven IBD research group (including Prof. Séverine Vermeire and other co-authors of TAGRID) write a yearly newsletter for all participants of their ongoing CCARE Biobank and related research. In this newsletter, the yearly progress is explained and elaborated future plans are discussed. Both researchers and clinicians actively participate in a yearly meeting organized for patients and their family members with the help of the Flemish Crohn’s and Colitis patient association (CCV). Articles are written regularly for the quarterly magazine of the CCV, in which the topic on results of this research project will also be distributed through all of these channels.

## EXTENDED DATA

**Figure 1.**
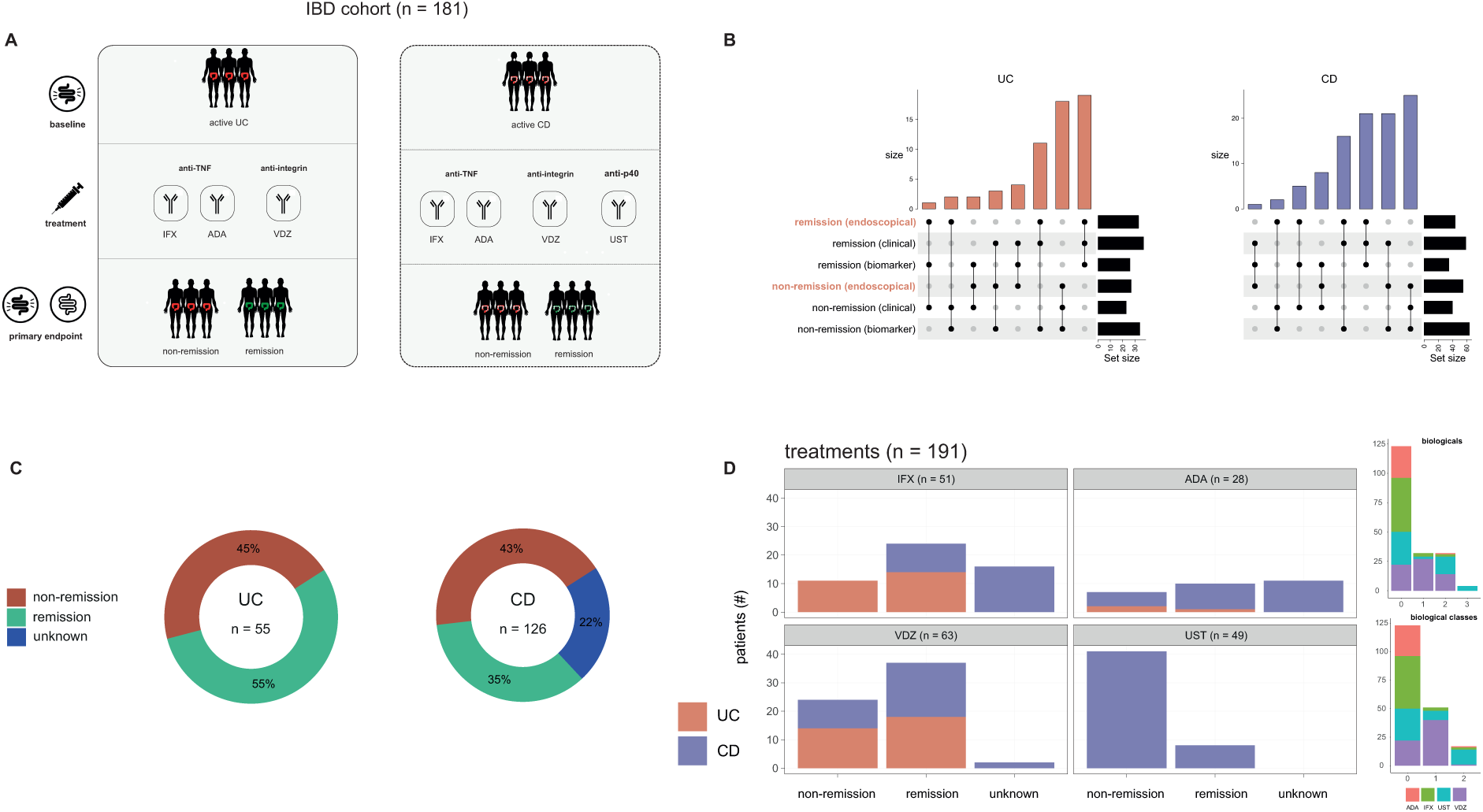
A prospective multi-therapeutic IBD cohort. **A**, Study design of a prospective active IBD cohort (*n=345*) undergoing biological treatment. Faecal samples (*n=432*) were selected from patients (*n=181*) based on availability and pairing. Patients had either active UC (*n=55*), or active CD (*n=126*) at baseline (pre-intervention). They were treated with one of four biologicals belonging to three treatment classes (anti-TNFs (IFX and ADA), anti-integrins (VDZ) or anti-p40 (UST)). At primary endpoint (post-intervention; w14 for UC; w24 for CD) patients were re-evaluated. **B**, SetUp plot shows overlap between post-intervention disease activity indices for IBD subtypes (remission and non-remission) as measured by different criteria (endoscopical, clinical and biomarker). **C**, Post-intervention disease activity for IBD subtypes, as measured by endoscopic evaluation (golden standard). **D (left)**, post-intervention disease activity for IBD subtypes shown per treatment as measured by endoscopic evaluation. **D (right)**, Treatment history of IBD patients shows number of biologicals (or biological classes) taken before. Abbreviations: Infliximab (IFX), Adalimumab (ADA), Vedolizumab (VDZ), Ustekinumab (UST), Ulcerative colitis (UC) and Crohn’s disease (CD), Inflammatory bowel disease (IBD).

**Figure 2.**
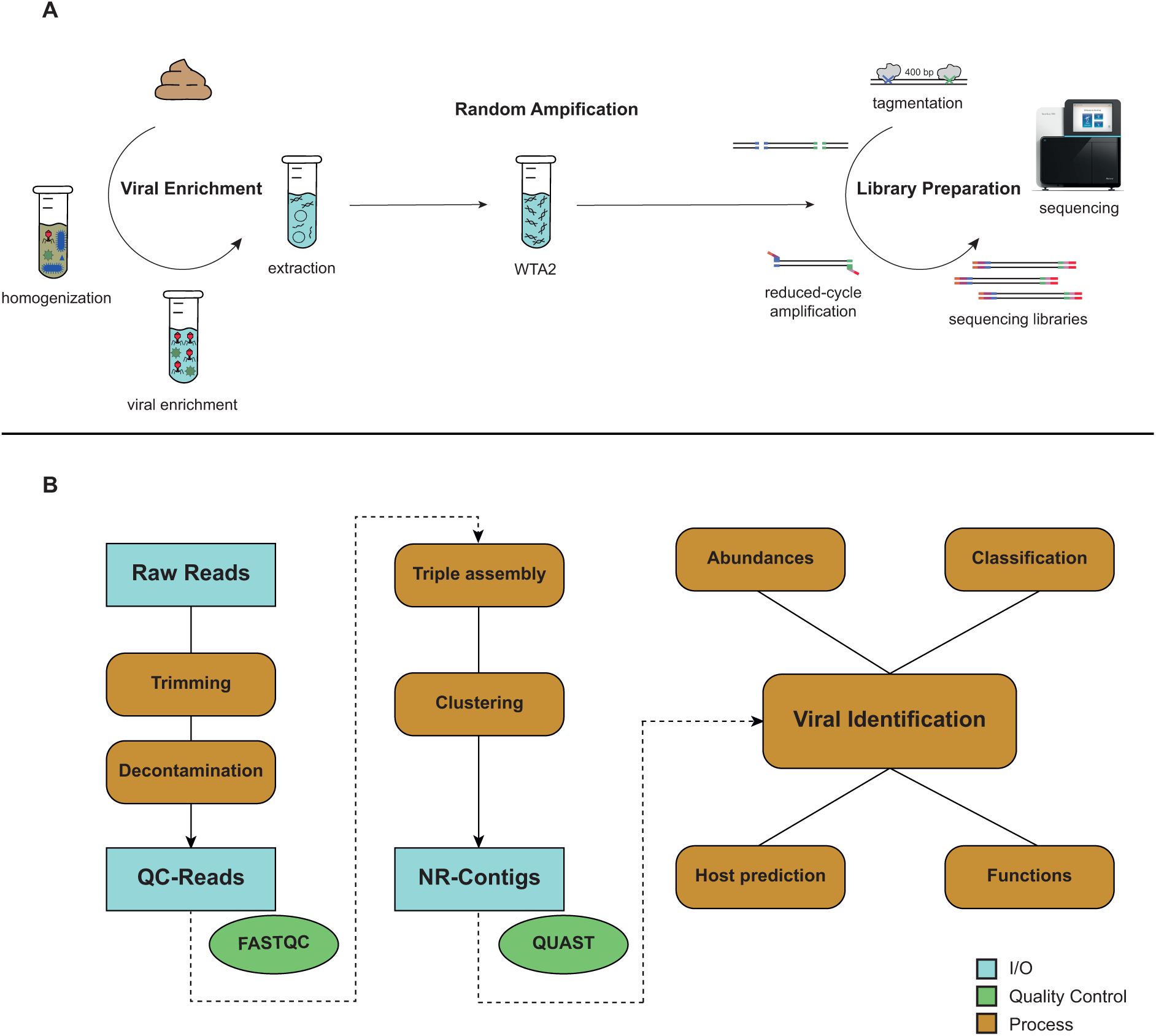
Viral metagenomic methods to analyse the gut virome. **A**, Overview of wetlab sample preparation using the NetoVIR protocol. The protocol was customized to enrich, reverse transcribe, random amplify and sequence viruses in a range of biological samples. **B**, Overview of drylab (bioinformatics) analyses of the gut virome. Raw reads are trimmed and decontaminated to obtain high-quality reads. The high-quality reads are *de novo* assembled with metaSPAdes into long contiguous sequences (contigs) using a three-step approach (see Methods for more details). Contigs are clustered to obtain a non-redundant contig set. Viruses are identified, classified and abundances are calculated. Functional characteristics of the identified viruses and their host are determined. Abbreviations: Input/Output (I/O), Contiguous sequences (contigs).

**Figure 3.**
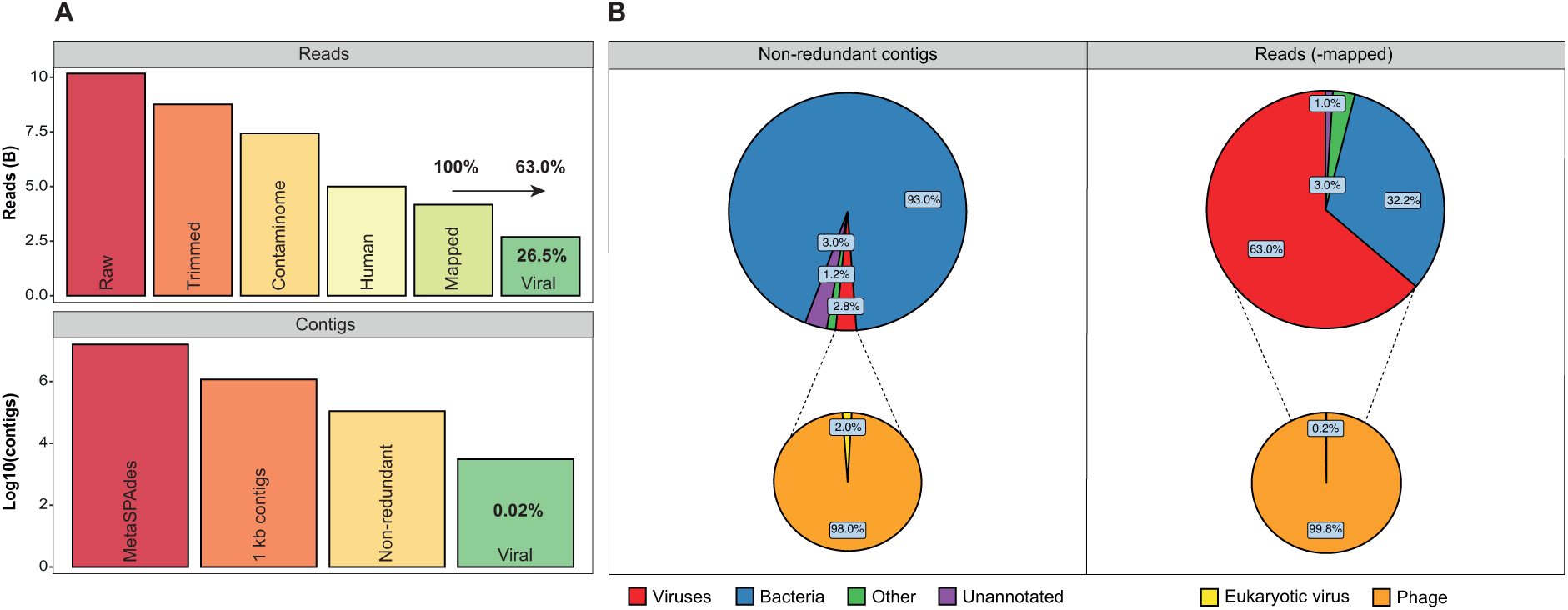
Quality steps and members of IBD viral metagenome. **A**, Reads and contigs per quality control step for the IBD shotgun dataset (raw reads=10.2 billion, metaSPAdes contigs=15.6 million). Viral contigs (0.02% of metaSPAdes contigs) represent non-redundant contigs identified as prokaryotic or eukaryotic viruses. Viral reads (26.5% of raw reads) represented quality-controlled reads mapped to non-redundant viral contigs (viral reads=2.7 billion, viral contigs=3033). **B**, Pie chart of non-redundant contigs showing distribution for each category (viruses, bacteria, Other (archaea, protozoa and eukaryota) and unannotated (dark matter)) and for the viral (eukaryotic viruses and phages) category (left). Pie chart of quality-controlled mapped reads showing distribution for each category and for the viral category (right).

**Figure 4.**
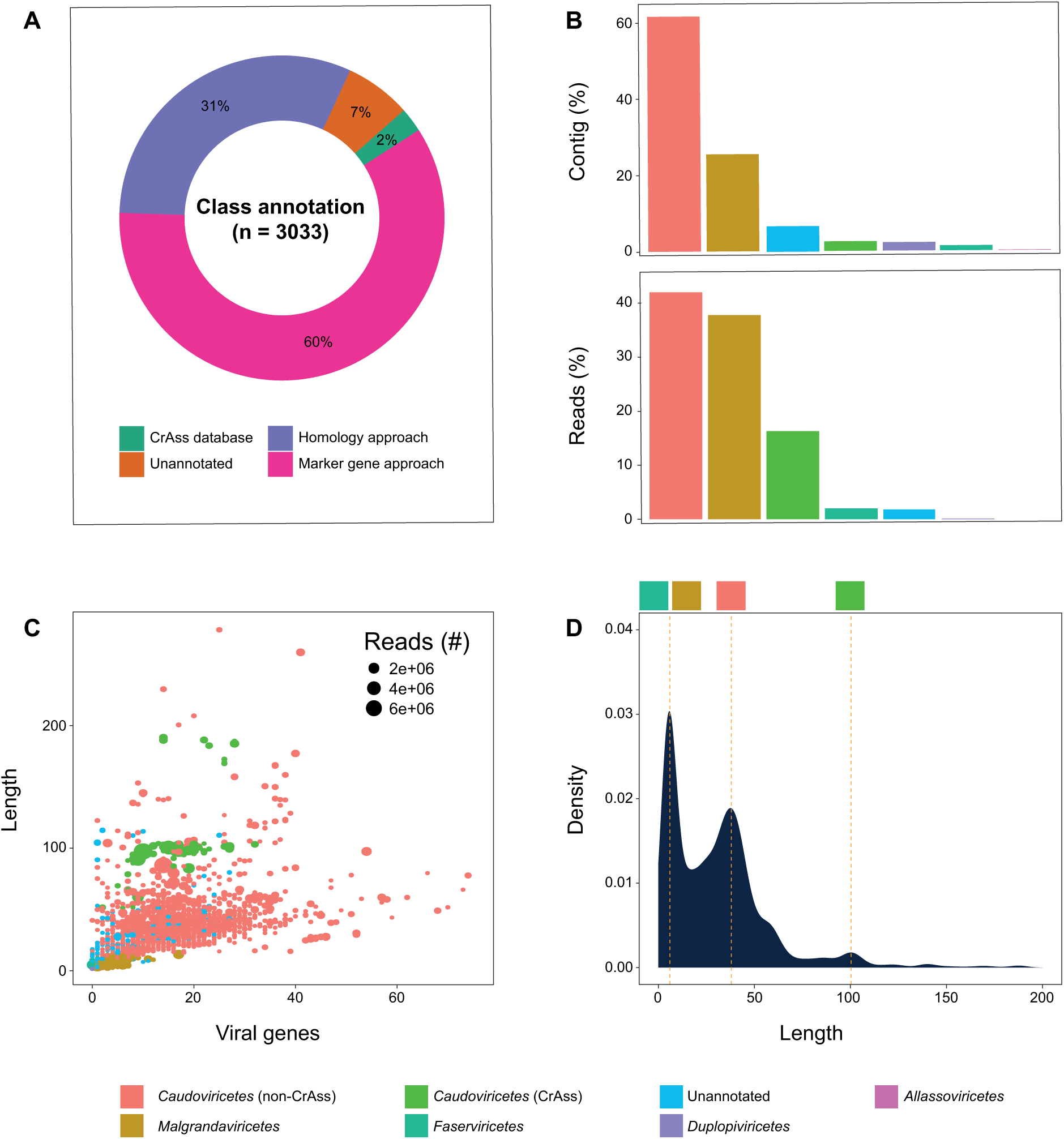
Viral classification by combining different approaches. **A**, Virome classification on class level taxonomy based on a combination of different approaches. First, CrAss-like phages (*Caudoviricetes* [CrAss]) are classified by comparing viral contigs (*n=3033*) to a custom-made CrAss phage database (2% of viral contig). Secondly, remaining viral contigs are classified based on a range of homology-based approaches such as Blastn, Diamond and CAT (lowest-common ancestor, 31% of viral contigs). Lastly, remaining viral contigs are classified based on a marker gene approach (60% of viral contigs) and a limit set of remained unclassified (7% of viral contigs). A total of 93% of the gut virome could be classified on class-level taxonomy. **B**, Phageome composition on class level in the prospective IBD cohort (contigs/reads). **C, D**, Scatterplot and density plot showing the class level classification over the length (maximum of 200kb) of the phage contig to estimate accuracy of classification (median length coloured by yellow dash line on D). The median of *Caudoviricetes* [CrAss] is 105.8 kb (literature:97kb), *Caudoviricetes* [non-CrAss] is 39.8 kb (literature:18kb-500kb), *Malgrandaviricetes* is 5.8 kb (literature:5.3kb-6.1kb) and *Faserviricetes* is 6.2 kb (literature:5.5kb-10.6kb). Genomes sizes (kb) of phages in literature were derived from International Committee for Taxonomy of Viruses (ICTV).

**Figure 5.**
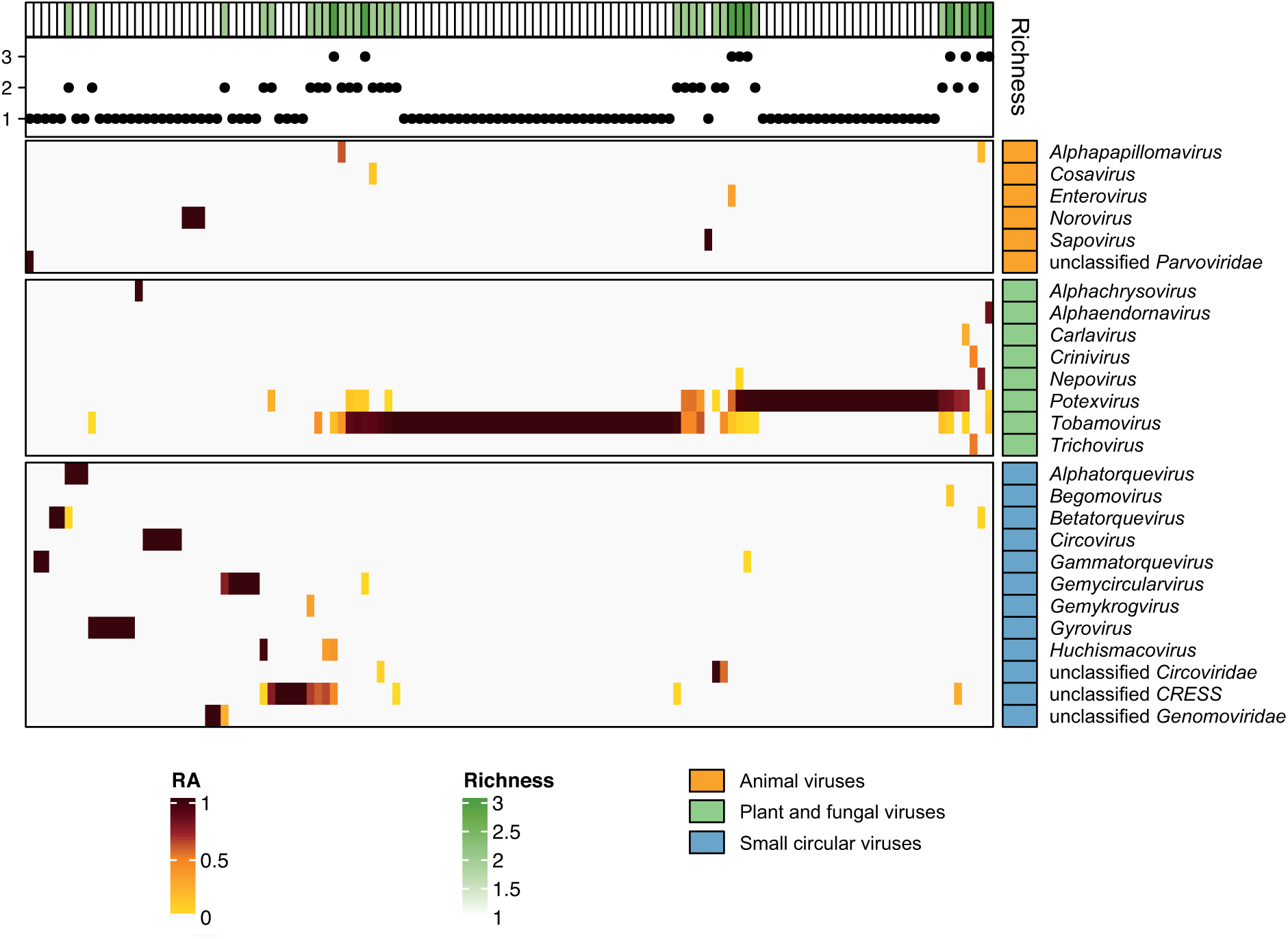
The eukaryotic virome of IBD patients undergoing biological therapy. A complex heatmap showing the relative abundance of the eukaryotic virome (genus level, *n=124*, eukaryotic viral presence=32.9%) in IBD patients. Eukaryotic viral genera are grouped based on a known (animal and plant/fungal viruses) or unknown (small circular viruses) host. Eukaryotic viruses were annotated based on homology-based classification (AS ≥ 0.1). Observed richness of eukaryotic viruses per sample (top). Abbreviations: Alignment score (AS).

**Figure 6.**
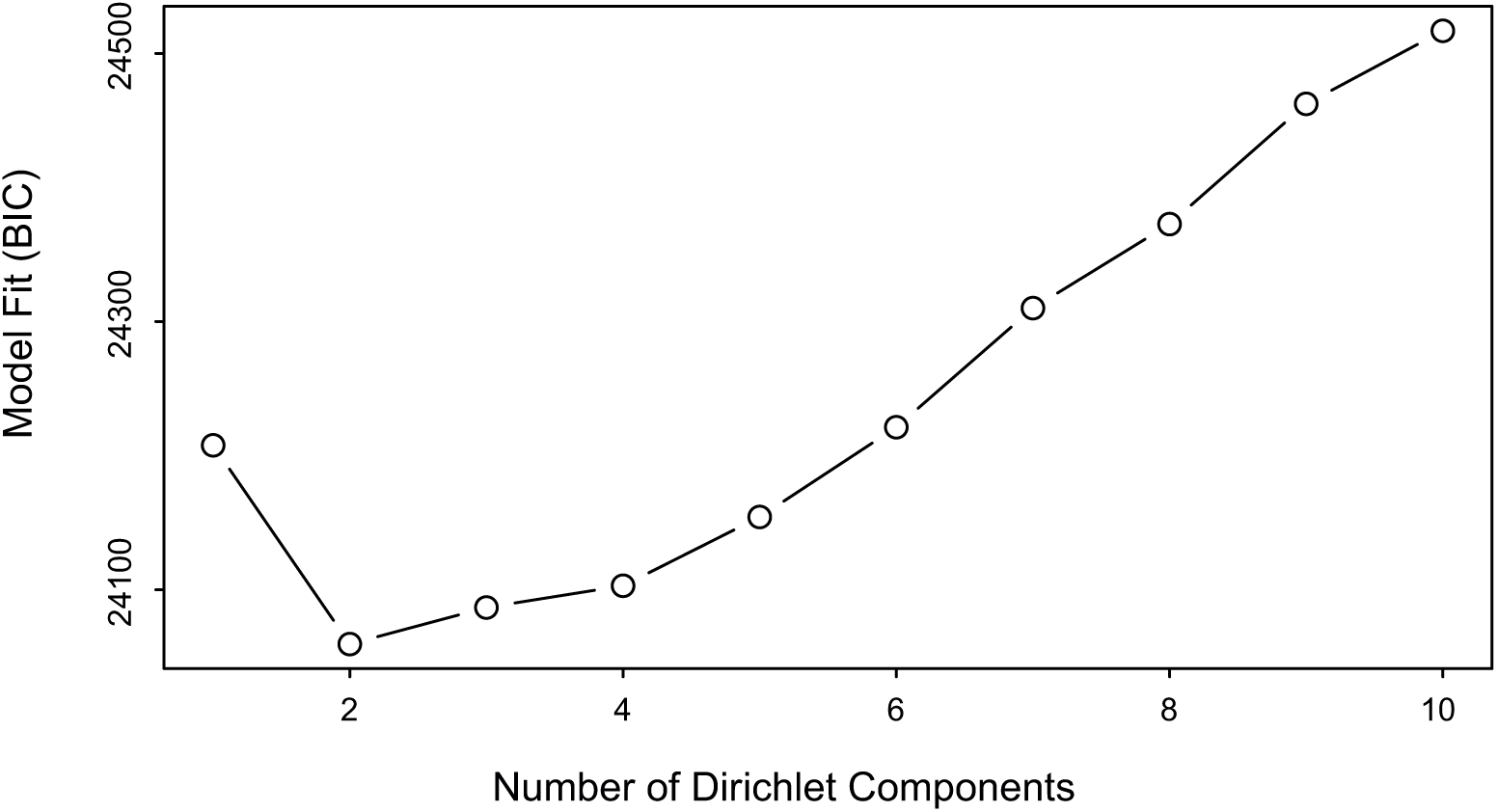
Identification of the optimal number of clusters for the DMM-based community typing of the IBD cohort. The optimal number of clusters (community types) is identified based on the Information criterion (minimum BIC) in the IBD cohort (*n=377*). The optimal number of clusters is two.

**Figure 7.**
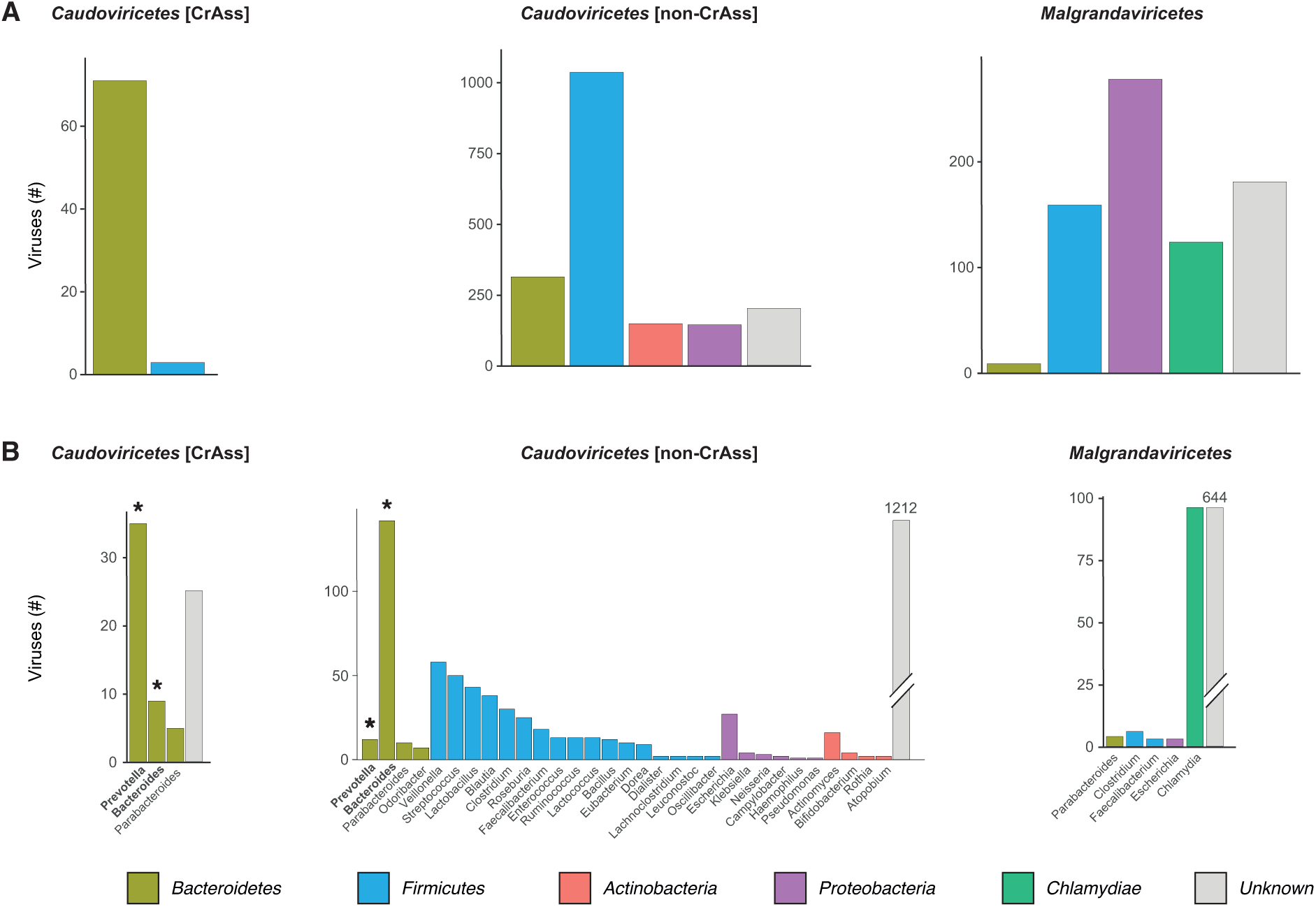
Bacterial hosts of major phages classes. **A**, Barplot showing predicted bacterial host phyla of the three major phage classes (>1% of reads, *Caudoviricetes* [CrAss], *Caudoviricetes* [non-CrAss] and *Malgrandaviricetes*). Host prediction for bacterial phyla could be made for 83.6% of the phages (16.4% phages with unknown host are coloured in gray). **B**, Barplot showing bacterial host genera of major phage classes (>1% of reads, *Caudoviricetes* [CrAss], *Caudoviricetes* [non-CrAss] and *Malgrandaviricetes*). The predicted bacterial hosts encompassing ≥1% of the reads are marked in bold with an astrix. Host prediction for bacterial genera could be made for 27.7% of the phages (72.3% phages with unknown host are coloured in gray). False positive host predictions are known to increase with shorter length of the phage genomes and therefore must be interpreted with caution (eg. *Malgrandaviricetes*).

**Figure 8.**
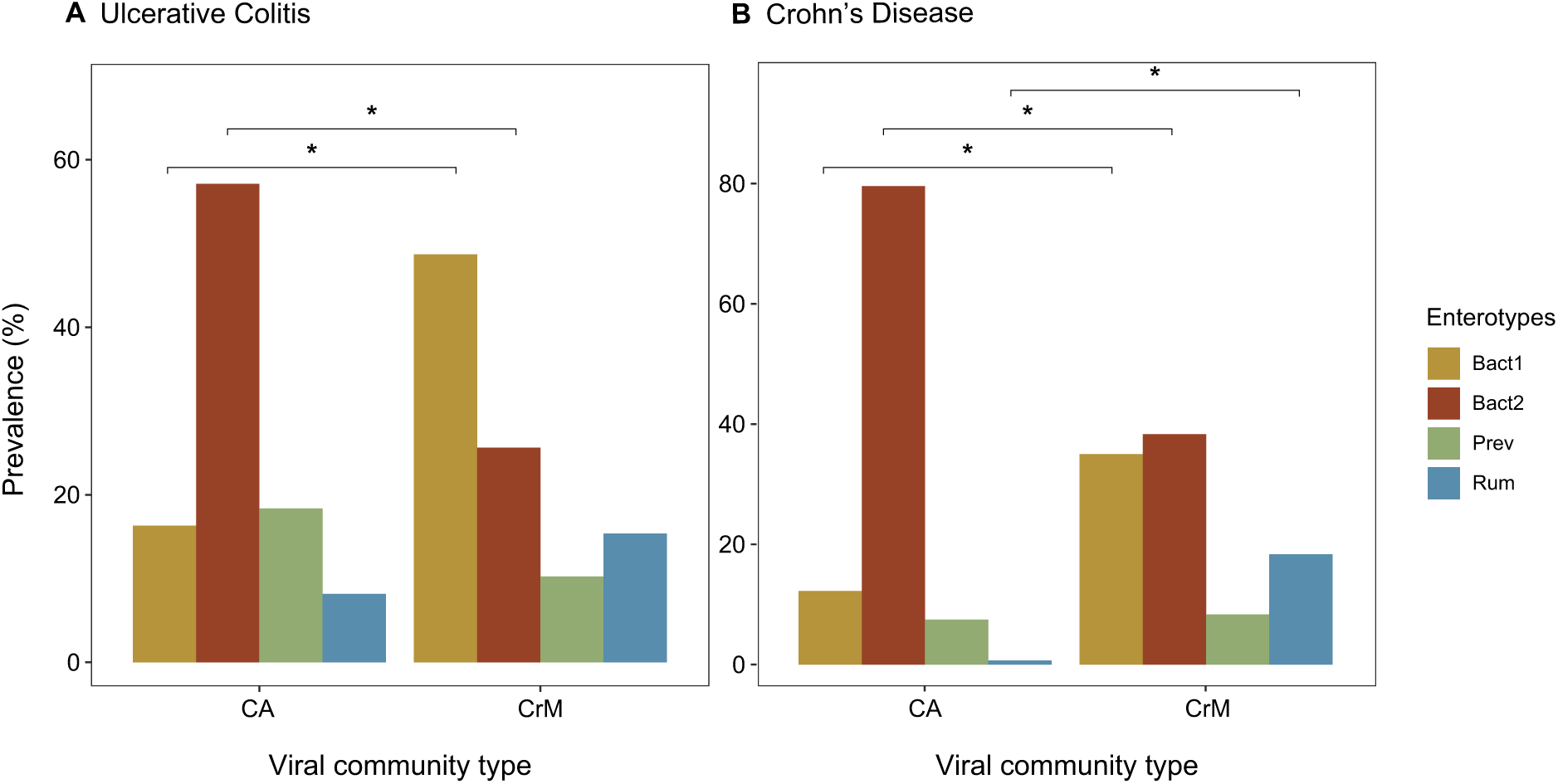
Enterotype distribution of the IBD cohort stratified according to viral community types. **A**, Enterotype (Bact1, Bact2, Prev and Rum) distribution stratified according to viral community types for ulcerative colitis patients (comparison Bact2, *n=88*, proportion test, AdjP<0.05). **B**, Enterotype distribution stratified according to viral community types for Crohn’s disease patients (comparison Bact2, *n=207*, proportion test, AdjP<0.05). Adjustment for multiple testing (AdjP) was performed using the Benjamini-Hochberg method. Significant associations (AdjP<0.05) were visualized by an astrix (*). Abbreviations: *Bacteroides* 1 (Bact1), *Bacteroides* 2 (Bact2), *Prevotella* (Prev) and *Ruminococcus* (Rum).

**Figure 9.**
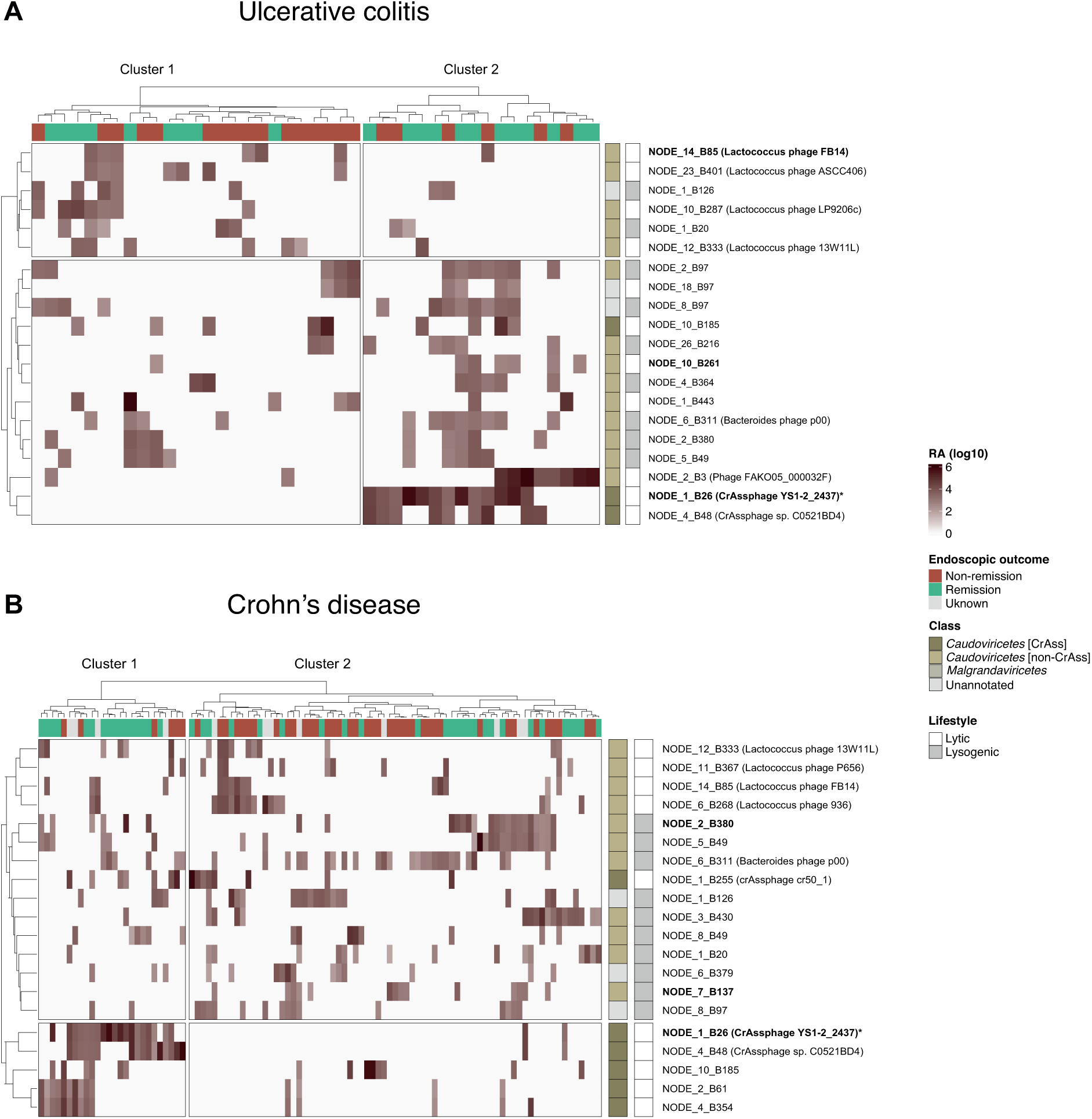
20 Most shared phages within the pre-interventional samples of the IBD cohort. **A,** Relative abundance complex heatmap of 20 most shared phages of baseline (pre-intervention) samples of UC patients (*n=44*). A total of 43/44 (97.7%) samples of UC patients contained at least 1 virus present at baseline. **B,** Relative abundance complex heatmap of 20 most shared phages of baseline (pre-intervention) samples of CD patients (*n=114*). A total of 99/114 (87.6%) samples of CD patients contained at least 1 virus present at baseline. Hierarchical clustering (euclidean with Ward.D) separating heatmap into 2 clusters (cluster 1 and cluster 2). Top annotation of the complex heatmap showing patients endoscopic outcome (non-remission, remission and unknowns). Right annotation showing phage class (*Caudoviricetes* [CrAss] and *Caudoviricetes* [non-CrAss], *Malgrandaviricetes* and unannotated), phage lifestyle (lytic and lysogenic) and phages names (NODES, shown if AS > 0.1). Names in bold represent phages that are differential abundant as well (Lefse analysis in main figure 5a). Abbreviations: Alignment score (AS), Ulcerative colitis (UC), Crohn’s disease (CD), Inflammatory bowel disease (IBD), linear discriminant analysis effect size (Lefse) and relative abundance (RA).

## REFERENCES

1. Wang B, Yao M, Lv L, Ling Z, Li L. The Human Microbiota in Health and Disease. Engineering. 2017 Feb 1;3(1):71–82.

2. Glassner KL, Abraham BP, Quigley EMM. The microbiome and inflammatory bowel disease. J Allergy Clin Immunol. 2020 Jan 1;145(1):16–27.

3. Öhman L, Lasson A, Strömbeck A, Isaksson S, Hesselmar M, Simrén M, et al. Fecal microbiota dynamics during disease activity and remission in newly diagnosed and established ulcerative colitis. Sci Reports 2021 111 [Internet]. 2021 Apr 21 [cited 2022 Feb 8];11(1):1–8. Available from: https://www.nature.com/articles/s41598-021-87973-7

4. Tedjo DI, Smolinska A, Savelkoul PH, Masclee AA, Van Schooten FJ, Pierik MJ, et al. The fecal microbiota as a biomarker for disease activity in Crohn’s disease. Sci Reports 2016 61 [Internet]. 2016 Oct 13 [cited 2022 Feb 8];6(1):1–10. Available from: https://www.nature.com/articles/srep35216

5. Prosberg M, Bendtsen F, Vind I, Petersen AM, Gluud LL. The association between the gut microbiota and the inflammatory bowel disease activity: a systematic review and meta-analysis. http://dx.doi.org/101080/0036552120161216587 [Internet]. 2016 Dec 1 [cited 2022 Feb 8];51(12):1407–15. Available from: https://www.tandfonline.com/doi/abs/10.1080/00365521.2016.1216587

6. Vieira-Silva S, Sabino J, Valles-Colomer M, Falony G, Kathagen G, Caenepeel C, et al. Quantitative microbiome profiling disentangles inflammation- and bile duct obstruction-associated microbiota alterations across PSC/IBD diagnoses. Nat Microbiol 2019 411 [Internet]. 2019 Jun 17 [cited 2022 Feb 8];4(11):1826–31. Available from: https://www.nature.com/articles/s41564-019-0483-9

7. Sokol H, Pigneur B, Watterlot L, Lakhdari O, Bermúdez-Humarán LG, Gratadoux JJ, et al. Faecalibacterium prausnitzii is an anti-inflammatory commensal bacterium identified by gut microbiota analysis of Crohn disease patients. Proc Natl Acad Sci [Internet]. 2008 Oct 28 [cited 2022 Feb 8];105(43):16731–6. Available from: https://www.pnas.org/content/105/43/16731

8. Costea PI, Hildebrand F, Manimozhiyan A, Bäckhed F, Blaser MJ, Bushman FD, et al. Enterotypes in the landscape of gut microbial community composition. Nat Microbiol 2017 31 [Internet]. 2017 Dec 18 [cited 2022 Jan 7];3(1):8–16. Available from: https://www.nature.com/articles/s41564-017-0072-8

9. Vandeputte D, Kathagen G, D’Hoe K, Vieira-Silva S, Valles-Colomer M, Sabino J, et al. Quantitative microbiome profiling links gut community variation to microbial load. Nat 2017 5517681 [Internet]. 2017 Nov 1 [cited 2022 Feb 8];551(7681):507–11. Available from: https://www.nature.com/articles/nature24460

10. Libby P. Statin drugs might boost healthy gut microbes. Nature. 2020 May 1;581(7808):263–4.

11. Clooney AG, Sutton TDS, Shkoporov AN, Holohan RK, Daly KM, O’Regan O, et al. Whole-Virome Analysis Sheds Light on Viral Dark Matter in Inflammatory Bowel Disease. Cell Host Microbe. 2019;26(6).

12. Zuo T, Lu X-J, Zhang Y, Cheung CP, Lam S, Zhang F, et al. Gut mucosal virome alterations in ulcerative colitis. Gut [Internet]. 2019 Jul 1 [cited 2019 Aug 17];68(7):1169–79. Available from: http://www.ncbi.nlm.nih.gov/pubmed/30842211

13. Norman JM, Handley SA, Baldridge MT, Droit L, Liu CY, Keller BC, et al. Disease-specific alterations in the enteric virome in inflammatory bowel disease. Cell. 2015;

14. Liang G, Cobián-Güemes AG, Albenberg L, Bushman F. The gut virome in inflammatory bowel diseases. Curr Opin Virol. 2021 Dec 1;51:190–8.

15. Zuo T, Lu XJ, Zhang Y, Cheung CP, Lam S, Zhang F, et al. Gut mucosal virome alterations in ulcerative colitis. Gut [Internet]. 2019 Jul 1 [cited 2022 Feb 8];68(7):1169–79. Available from: https://gut.bmj.com/content/68/7/1169

16. Liang G, Conrad MA, Kelsen JR, Kessler LR, Breton J, Albenberg LG, et al. Dynamics of the Stool Virome in Very Early-Onset Inflammatory Bowel Disease. J Crohn’s Colitis [Internet]. 2020 Nov 7 [cited 2022 Jun 17];14(11):1600–10. Available from: https://academic.oup.com/ecco-jcc/article/14/11/1600/5837025

17. Pérez-Brocal V, García-López R, Vázquez-Castellanos JF, Nos P, Beltrán B, Latorre A, et al. Study of the viral and microbial communities associated with Crohn’s disease: A metagenomic approach. Clin Transl Gastroenterol [Internet]. 2013 [cited 2022 Jun 17];4. Available from: https://journals.lww.com/ctg/Fulltext/2013/06000/Study_of_the_Viral_and_Microbial_Communities.2.aspx

18. Fernandes MA, Verstraete SG, Phan T, Deng X, Stekol E, Lamere B, et al. Enteric Virome and Bacterial Microbiota in Children with Ulcerative Colitis and Crohn Disease. J Pediatr Gastroenterol Nutr [Internet]. 2019 [cited 2022 Jun 17];68(1):30–6. Available from: https://journals.lww.com/jpgn/Fulltext/2019/01000/Enteric_Virome_and_Bacterial_Microbiota_in.8.aspx

19. Yutin N, Benler S, Shmakov SA, Wolf YI, Tolstoy I, Rayko M, et al. Analysis of metagenome-assembled viral genomes from the human gut reveals diverse putative CrAss-like phages with unique genomic features. Nat Commun [Internet]. 2021 Dec 1 [cited 2021 Jul 1];12(1):1–11. Available from: https://www.nature.com/articles/s41467-021-21350-w

20. Gulyaeva A, Garmaeva S, Ruigrok RAAA, Wang D, Riksen NP, Netea MG, et al. Discovery, diversity, and functional associations of crAss-like phages in human gut metagenomes from four Dutch cohorts. Cell Rep. 2022 Jan 11;38(2):110204.

21. Shkoporov AN, Khokhlova E V., Stephens N, Hueston C, Seymour S, Hryckowian AJ, et al. Long-term persistence of crAss-like phage crAss001 is associated with phase variation in Bacteroides intestinalis. BMC Biol [Internet]. 2021 Dec 1 [cited 2022 Feb 8];19(1):1–16. Available from: https://bmcbiol.biomedcentral.com/articles/10.1186/s12915-021-01084-3

22. Caenepeel C, Falony G, Machiels K, Verstockt B, Ferrante M, Sabino J, et al. Dysbiosis-associated stool features improve prediction of response to biological therapy (anti-TNF, alpha, anti-integrin and anti-interleukin 12/23) in inflammatory bowel disease. Under Revis Gastroenterol. 2022;

23. Schwarz D, Beuch U, Bandte M, Fakhro A, Büttner C, Obermeier C. Spread and interaction of Pepino mosaic virus (PepMV) and Pythium aphanidermatum in a closed nutrient solution recirculation system: effects on tomato growth and yield. Plant Pathol [Internet]. 2010 Jun 1 [cited 2022 Jun 18];59(3):443–52. Available from: https://onlinelibrary.wiley.com/doi/full/10.1111/j.1365-3059.2009.02229.x

24. Court DL, Oppenheim AB, Adhya SL. A new look at bacteriophage λ genetic networks. J Bacteriol [Internet]. 2007 Jan [cited 2022 Jul 4];189(2):298–304. Available from: https://journals.asm.org/doi/full/10.1128/JB.01215-06

25. Loddo I, Romano C. Inflammatory bowel disease: Genetics, epigenetics, and pathogenesis. Front Immunol. 2015;6(NOV):551.

26. Bassotti G, Antonelli E, Villanacci V, Nascimbeni R, Dore MP, Pes GM, et al. Abnormal gut motility in inflammatory bowel disease: an update. Tech Coloproctol [Internet]. 2020 Apr 1 [cited 2022 May 20];24(4):275–82. Available from: https://link.springer.com/article/10.1007/s10151-020-02168-y

27. Zuppi M, Hendrickson HL, O’Sullivan JM, Vatanen T. Phages in the Gut Ecosystem. Front Cell Infect Microbiol. 2022 Jan 4;11:1348.

28. Conceição-Neto N, Zeller M, Lefrère H, De Bruyn P, Beller L, Deboutte W, et al. Modular approach to customise sample preparation procedures for viral metagenomics: a reproducible protocol for virome analysis. Sci Rep. 2015;

