## Extended Data Methods for "Community types of the human gut virome are associated with endoscopic outcome in ulcerative colitis"

#### Ethical approval

The study was approved by the ethical commission of UZ Leuven (KU Leuven, reference number: S53684). Participants provided signed informed consent to participate with the study. The design of the study was in accordance with the Declaration of Helsinki and Belgian privacy law.

#### Study design

The prospective study consisted of 345 active IBD (239 CD and 106 UC) patients initiating biological therapy. They were included after attending the outpatient clinic of the UZ Leuven (Leuven, Belgium) following informed consent. IBD patients were diagnosed based on European Crohn and colitis organisation (ECCO) guidelines[1,2]. Active disease before patients were treated (baseline) met the criteria for initiating every biological therapy based on endoscopic evaluation. Patients were treated with anti-tumor necrosis factor (aTNF, 159 patients), vedolizumab (VDZ, 148 patients) and/or ustekinumab (UST, 78 patients). Disease activity for UC patients was based on patient report outcome 2 (PRO2) combining stool frequency and rectal bleeding, Mayo endoscopy subscore and faecal calprotectin[3]. Disease activity for CD patients was based on PRO2 combining stool frequency and abdominal pain, simple endoscopic score for Crohn's disease (SES-CD) or endoscopic absence of ulceration and faecal calprotectin. In addition, albumin, serum CRP and hemoglobin were used to establish disease activity as well. Therapeutic outcome to biological therapy was determined at week 14 (w14) for UC and week 24 (w24) for CD patients (primary endpoint). Therapeutic outcome was defined by endoscopic, clinical and biomarker remission of the patient. Endoscopic remission in UC patients was defined as Mayo endoscopy subscore ≤ 1 and in CD patients as SES-CD ≤ 3 and/or absence of ulceration. Clinical remission in UC patients was defined as PRO2 with no rectal bleeding and stool frequency ≤ 1. Clinical remission in CD patients was defined as PRO2 ≤ 7 (calculated as stool frequency x 2 + abdominal pain x 5). Biomarker remission in all patients was defined as faecal calprotectin < 150 mg/kg.

**Metadata collection**

The patients metadata consisted of anthropometrics, clinical parameters, and characteristics of faecal samples and are available in Supplementary Table 1. Anthropometric quantitative measurements are comprised of age (years), gender (male/female), smoking status (active smoker or non-smoker), BMI ($kg/m^2$), disease duration (years), Diagnosis (UC/CD) and biological history (number of therapies before). Clinical parameters consisted of disease location (Montreal classification and grouping UC with colonic CD patients in L2), serum CRP (mg/L), albumin (g/L), hemoglobin (g/dL), endoscopic and total mayo-score, SES-CD and PRO2. Faecal samples were characterized by assessing faecal calprotectin (mg/L), moisture content (%) and bacterial cell count. Faecal calprotectin concentrations were measured on frozen samples using the fCAL ELISA kit (Bühlmann, Schönenbuch, Switzerland). Faecal moisture was calculated after lyophilization as the percentage of mass loss from ± 0.2 g frozen aliquots (-80°C). Microbial loads were determined in ± 0.2 g frozen (-80°C) aliquots as described before[4,5].

**Sample selection**

A subset of faecal samples was collected from the original prospective study cohort (*n=345)* based on sample availability and pairing. Faecal samples (*n=432*) were collected from a total of 181 IBD (126 CD and 55 UC) patients. All IBD patients had active disease at baseline and were treated with one of four biological therapies (51 IFX, 28 ADA, 63 VDZ and 49 UST). Every patient comprised of a baseline and primary endpoint faecal sample, and 50 CD patients contained an additional w14 timepoint. Some patients (*n=10*) received 2 biological treatments. In this study, the term (non-)remission was used to refer to samples of (non-)responding patients based on endoscopic outcome, irrespective of the timepoint of sample collection (baseline/primary endpoint).

**Viral metagenomics**

The NetoVIR protocol was used to prepare faecal samples for viral metagenomics as described before (Extended Data Figure 2A)[6]. Faecal samples were aliquoted in capped O-ring tubes (50-100 mg) and suspended in sterile dPBS (10%). The suspension was homogenized using the PRECELLYS homogenizer (Bertin Technologies) for 1 min at 3,000 rpm following a centrifugation step for 3 min at 17,000 g. The supernatant was filtered through a 0.8 µm PES filter (Sartorius) and a nuclease treatment was performed using a combination of benzonase (Novagen) and micrococcal nuclease (New England Biolabs) at 37°C for 2h. Extraction of viral nuclei acids was performed without addition of carrier RNA using the QIAMP® Viral RNA mini kit (60µL, Qiagen, Venlo, Netherlands). Nucleic acids were randomly amplified using the Complete Whole Transcriptome Amplification kit (WTA2, Sigma-Aldrich) with small adaptations (94°C for 2 min, and 17 cycles of 94°C for 30 sec and 70°C for 5 min). The amplified PCR product was purified using the MSB Spin PCRapace kit (Invitek Molecular) and concentration was measured using the Qubit™ dsDNA HS Assay Kit. The preparation of the sequencing libraries was performed using the Nextera XT DNA Library kit (Illumina) and purified with Agencourt AMPure XP beads (Beckman Coulter). Quality and library sizes were evaluated using a High Sensitivity DNA kit on the Bioanalyzer 2100 (Agilent Technologies). Sequencing of the libraries was carried out by VIB Nucleomics Core in 2 runs on an Illumina NovaSeq6000 S2 sequencer (2x150bp, paired end).

### Bioinformatic analyses

Computational analyses on an input of 10.2 billion paired end reads (1.52 TB) was carried out as described before (Extended Data Figure 2B)[7]. The average was 23.6 (range 7.09 to 50.3) million reads per sample. Raw reads were trimmed to remove low-quality bases, ambiguous bases and adapter sequences using Trimmomatic v0.39[8]. Trimmed reads were decontaminated upon alignment to the reference human genome (hg38, BioProject=PRJNA31257), and contaminome sequences present in negative controls with bwa-mem2 v2.0[9]. The average number of quality-controlled reads per sample was 11.6 (range 6,669 to 33.3) million per sample. These reads were *de novo* assembled in a set of long contiguous sequences (contigs) using MetaSPAdes v3.15.1 with k-mer length of 21, 33, 55 and 77[10]. To reduce fragmentation of contigs with a very high coverage the assembly step was repeated two more times with respectively 10% and 1% of the reads (triple assembly). Per sample, the contigs of the 3 assemblies were merged, clustered and contigs with a sequence length lower than 1,000 bp were discarded. A second clustering was performed, this time across all samples to remove redundancy and obtain a set of non-redundant (NR) contigs at 95% ANI and 80% coverage using CheckV's clustering scripts[11]. Abundances were calculated per sample by mapping quality-controlled reads to the set of NR contigs using bwa-mem2 v2.0, provided that the respective sample had a member in the cluster of contigs that NR contigs represent[9]. To avoid false positive identification of phages in a sample, NR contigs with a horizontal coverage of 70% or lower were discarded. To correct for sequencing depth viral metagenomes were subsampled (rarefied) to ± 1 million reads (*n=377*) thereby discarding 55 samples.

### Eukaryotic viruses

Identification and classification (genus/family) of eukaryotic viruses was performed using well-annotated public databases. The homology-based approaches compared the

NR-contig set against a NR protein sequence database (Jan 8, 2021) using DIAMOND v0.9.30.131 (sensitive mode) and CAT v4.6, and against the NCBI nucleotide database (April 18, 2021) using BLASTN v2.7.1 (e-value ≤ 1e-10)[12–14]. Classification was derived from the principle of lowest common ancestor as determined by ktClassifyBLAST module in KronaTools v2.8[15].

**Prokaryotic viruses**

Identification of prokaryotic viruses (bacteriophages/phages) was performed using VirSorter2 v2.2.3 (--min-score ≥ 0.5)[16]. Genome completeness of the NR-contigs was determined with CheckV v0.5.1[11]. Bacteriophages identified with VirSorter2 and an adequate quality tier (≥ 50% completeness) were selected for further analyses. Classification was performed by a combination of homology-based approaches and a marker gene approach. Homology-based approaches were described in the previous section for eukaryotic viruses. Additionally, phage classification was expanded by marker gene approaches using Cenote-Taker2 v2.0.1 (Extended Data Figure 3)[17]. Early phage taxonomy has been largely based upon phage morphology. The International Committee on Taxonomy of Viruses (ICTV) has undertaken large efforts to reorganize viral taxonomy based on genetic similarities in an ongoing challenge to optimize phage taxonomy[18,19]. Although great progress has been made, linking current taxonomies to sequencing data at family (or any other lower) taxonomical rank remains inadequate. To minimize false annotations, phage classification was specified on the class taxon.

The lifestyle of bacteriophages was determined based on the appearance of lysogeny-specific genes. These genes were predicted using the functional annotation module of Cenote-Taker2 and can be found in Supplementary Table 6[14]. The functional annotation module of Cenote-Taker2 was also used to screen for toxins and reverse transcriptase

(RT) characteristics. The bacterial host of phage genomes was predicted using Random Forest Assignments of Hosts (RaFAH) v0.3[20]. Bacterial hosts were predicted on the phyla taxon (--min_cutoff ≥ 0.14) and the genus taxon (--min_cutoff ≥ 0.50) as suggested by the authors.

**CrAss-like bacteriophages**

A custom database of 998 CrAss-like bacteriophages was created by combining the CrAss-like genomes of 3 large datasets. A total of 55 genomes were found in RefSeq, 249 genomes in *Guerin et al*[16], and 694 genomes in *Yutin et al*[17]. Bacteriophages selected in previous section were compared against this custom nucleotide database using BLASTN (e-value ≤ 1e-5, %cov ≥ 10,000 bp) to identify CrAss-like viruses[9]. A total of 74 CrAss-like viruses (2.4%) were identified. accounting for 16.2% of the viral reads (Supplementary Table 2). CrAss-like viruses were classified as a separate group within the class taxon of *Caudoviricetes* and named Caudoviricetes [CrAss].

**Viral community typing**

Bacterial community-typing ('enterotyping') is based on Dirichlet Multinomial Mixture (DMM) modelling and allows for the stratification of individuals based on their gut microbiome[6,21,22]. Briefly, DMM includes a probabilistic modelling that groups samples from the same community, and thereby reproducibly identifies microbiome configurations without making any assumptions regarding the putative discrete nature of the strata[,21,22]. A viral counterpart of these enterotypes ('viral community typing) might allow researchers to stratify individuals based on their gut virome. In addition, viral community types could be associated to covariates, as mentioned before[21]. A prerequisite for viral community typing is a high degree of shared viral groups between individuals which is problematic due to a high virome individuality[23]. Homology-based approaches to obtain reliable lower ranks (e.g. genera) are currently unreliable[18]. For that reason, in this study viral groups that clustered together based on amino acid similarity were assigned to the same genus-like groups, as described in *Nayfach et al*[24]. A total of 874 viral genus-like groups were detected within this IBD cohort (median per sample=26). Viral community typing of the genus-like groups (rarefied) abundances (≥20% prevalence) with logarithmic transformation was performed based on the Dirichlet multinomial mixtures approach as provided in the DirichletMultinomial R package[25]. The number of mixture components was determined to be two according to the Bayesian information criterion (BIC) score (*n=363*, Extended Data Figure 6). The mean probability for cluster assignment was 0.93 (median=1, Supplementary Table 4).

**Diversity, abundances and virome compositional visualization**

Alpha-diversity indices (observed richness and Shannon diversity) were calculated on the abundance table using phyloseq[26]. Differential abundance analysis was used to identify features whose relative abundances differ between two or more groups of samples by linear discriminant analysis effect size (LEfSe) as implemented in the microbiomeMarker R package[27]. Virome inter-individual variation was calculated using Bray-Curtis dissimilarity on the genus-like groups abundances with logarithmic transformation and visualized by principal coordinate analysis (PCoA). Additionally, enterotyping (or bacterial community typing) of the genus-level abundance microbial profiles were performed as part of another paper[28].

**Explanatory analyses of virome compositional variation**

The contribution of metadata to the virome variation (genus level, Bray-Curtis) was determined using univariate distance-based redundancy analysis (dbRDA) using the *capscale* function as described in the vegan R package[29]. The contribution of significant metadata variables on the first two principal coordinates were determined using the *envfit* function as implemented in the vegan package (univariate dbRDA) and were plotted as arrows on the PCoA plot[29]. The non-redundant cumulative contribution of metadata to the virome variation with maximum explanatory power (genus level, Bray-Curtis) was determined using single multivariate dbRDA by forward model selection with the *ordiR2step* function in vegan. Metadata variables were included in multivariate analyses if they showed a significant contribution to virome variation in previous univariate dbRDA. To assess the contribution of a variable on the virome variation between timepoints a subset of samples was assessed (limited to baseline and primary endpoint samples) by pairing the analysis using the *capscale* function and conditioning on patient identifier (paired dbRDA).

**Metadata association to community types**

Logistic regression (Logit) allows for the association between metadata and community types. Simple logistic regression modelled the association between a binomial response variable (community type CrM presence (true/false)) using one non-redundant explanatory variable. The Logit link function was applied (generalized linear model with binomial link function), as provided by the glm function in the stats package.

**Prevalence, quantification, and classification of potentially predictive biomarkers**

Viral extracts (60 μL) of pre-intervention samples (without VLP enrichment) were used to determine the prevalence and amount of previously identified phages with predictive potential using qPCR (Supplementary Table 25). To evaluate the novelty of the phages compared to current databases, blastn alignment score (AS) was given by multiplying average nucleotide identity (ANI) with the alignment coverage (AC). Phages were given a species classification based on the closest match if AS > 0.1 or were classified as unannotated if AS < 0.1. In total 5 novel phage species were identified to potentially have a predictive capacity with a poor match to known phages, and therefore have been given a novel name. The discovered phages were named CrAssella-R (NODE_1_B26, species = CrAssphage YS1-2_2437, AS = 0.38), Croides-R (NODE_2_B380), Croccus-NR (NODE_14_B85, species = Lactococcus phage FB14, AS = 0.41), Croides-NR (NODE_7_B137) and Cripes-R (NODE_10_B261). To evaluate the prevalence and amount, primers, probes, and standards specific to each phage were designed based on the alignment of medium to-complete genomes as determined by CheckV (Supplementary Table 26). Quantification of the viral load in each pre-intervention sample was performed for each phage in duplicate qPCRs. The qPCR mix was made by adding 5 µL viral extract to a mastermix of 15 µL. The mastermix was composed of 5 µL TaqMan Fast Virus 1-Step Mastermix (ThermoFisher), 5 µL sterile water, 1 µL probes (5µM) and 2 µL forward/reverse primer (10µM). The standards contained a known concentration of oligonucleotides and were used to establish a calibration curve by serial dilution ($10^{11}$ to $10^{3}$ viral copies). This calibration curve was used for calculating the concentration of the respective phages in the pre-intervention samples. The total viral copies of each sample were calculated by multiplying the qPCR results by a dilution factor of 12 (dilution factor: 5 µL out of 60 µL viral extract). The limit of detection to determine phage marker prevalence was set at 100 viral copies per mL. The limit of quantification to determine phage marker concentration was set at 500 viral copies per mL.

#### Predictive model for therapeutic outcome

Phage marker concentration for each pre-intervention samples was used to calculate the IBD predictive value. The predictive value of IBD phage markers was calculated by $log10(\frac{B26+B261+1}{B85+1})$ and $log10\left(\frac{B26+B380+1}{B137+1}\right)$ for UC and CD patients, respectively. Simple logistic regression modelled the association between a binomial response variable (endoscopic remission (true/false)) using the IBD predictive value (no prediction excluded). The Logit function was applied (generalized linear model with binomial link function), as provided by the glm function in the stats package. At last, a ROC (receiver operating characteristic) curve was calculated illustrating the predictive ability of the phage markers using the pROC package.

**Statistical analyses**

Statistics were performed in R using the packages phyloseq, DirichletMultinomial, vegan, robustrank and stats packages[25,26,29–31]. All statistical tests were non-parametric, two-sided, and significance was defined as P < 0.05. Multiple testing correction was applied where appropriate using Benjamini-Hochberg (BH) method, and significance was defined as AdjP < 0.05. Wilcoxon effect size was calculated by $r = Z/\sqrt{N}$ and Chi-squared effect size was calculated by $r = \sqrt{\chi^2}/N$, as implemented in the rstatix package.

**Data availability**

Metadata can be found in Supplementary Table 1. The raw sequence data were deposited to the NCBI Sequence Read Archive under the BioProject accession number PRJNA804384. Sequences (predictive markers) were deposited to GenBank under the following accession numbers: ON493177-ON493181.

**Code availability**

The ViPER (Virome Paired-End Reads pipeline) script was used to process raw paired-end reads and is publicly available at **https://github.com/Matthijnssenslab/ViPER**. All the data required to reproduce virome analyses will be made available at https://github.com/Matthijnssenslab/IBDVirome.

**REFERENCES**


1. Harbord M, Eliakim R, Bettenworth D, Karmiris K, Katsanos K, Kopylov U, et al. Third European Evidence-based Consensus on Diagnosis and Management of Ulcerative Colitis. Part 2: Current Management. J Crohn's Colitis [Internet]. 2017 Jul 1 [cited 2021 Feb 14];11(7):769–84. Available from: https://academic.oup.com/ecco-jcc/article/11/7/769/2962457
2. Van Assche G, Dignass A, Panes J, Beaugerie L, Karagiannis J, Allez M, et al. The second European evidence-based consensus on the diagnosis and management of Crohn's disease: Definitions and diagnosis [Internet]. Vol. 4, Journal of Crohn's and Colitis. Oxford Academic; 2010 [cited 2021 Feb 13]. p. 7–27. Available from: https://academic.oup.com/ecco-jcc/article/4/1/7/402141
3. Loddo I, Romano C. Inflammatory bowel disease: Genetics, epigenetics, and pathogenesis. Front Immunol. 2015;6(NOV):551.
4. Vieira-Silva S, Sabino J, Valles-Colomer M, Falony G, Kathagen G, Caenepeel C, et al. Quantitative microbiome profiling disentangles inflammation- and bile duct obstruction-associated microbiota alterations across PSC/IBD diagnoses. Nat Microbiol 2019 411 [Internet]. 2019 Jun 17 [cited 2022 Feb 8];4(11):1826–31. Available from: https://www.nature.com/articles/s41564-019-0483-9
5. Sokol H, Pigneur B, Watterlot L, Lakhdari O, Bermúdez-Humarán LG, Gratadoux JJ, et al. Faecalibacterium prausnitzii is an anti-inflammatory commensal bacterium identified by gut microbiota analysis of Crohn disease patients. Proc Natl Acad Sci [Internet]. 2008 Oct 28 [cited 2022 Feb 8];105(43):16731–6. Available from:

https://www.pnas.org/content/105/43/16731

6. Costea PI, Hildebrand F, Manimozhiyan A, Bäckhed F, Blaser MJ, Bushman FD, et al. Enterotypes in the landscape of gut microbial community composition. Nat Microbiol 2017 31 [Internet]. 2017 Dec 18 [cited 2022 Jan 7];3(1):8–16. Available from: https://www.nature.com/articles/s41564-017-0072-8
7. Simsek C, Corman VM, Everling HU, Lukashev AN, Rasche A, Maganga GD, et al. At least seven distinct rotavirus genotype constellations in bats with evidence of reassortment and zoonotic transmissions. MBio. 2021;12(1).
8. Bolger AM, Lohse M, Usadel B. Trimmomatic: A flexible trimmer for Illumina sequence data. Bioinformatics. 2014;
9. Md V, Misra S, Li H, Aluru S. Efficient architecture-aware acceleration of BWA-MEM for multicore systems. In: Proceedings - 2019 IEEE 33rd International Parallel and Distributed Processing Symposium, IPDPS 2019. Institute of Electrical and Electronics Engineers Inc.; 2019. p. 314–24.
10. Bankevich A, Nurk S, Antipov D, Gurevich AA, Dvorkin M, Kulikov AS, et al. SPAdes: A New Genome Assembly Algorithm and Its Applications to Single-Cell Sequencing. J Comput Biol. 2012;
11. Nayfach S, Camargo AP, Schulz F, Eloe-Fadrosh E, Roux S, Kyrpides NC. CheckV assesses the quality and completeness of metagenome-assembled viral genomes. Nat Biotechnol 2020 395 [Internet]. 2020 Dec 21 [cited 2022 Jan 7];39(5):578–85. Available from: https://www.nature.com/articles/s41587-020-00774-7
12. Buchfink B, Xie C, Huson DH. Fast and sensitive protein alignment using DIAMOND. Nat Methods [Internet]. 2015 Jan 17 [cited 2019 Aug 26];12(1):59–60. Available from: http://www.nature.com/articles/nmeth.3176
13. Altschul SF, Gish W, Miller W, Myers EW, Lipman DJ. Basic local alignment search

tool. J Mol Biol [Internet]. 1990 Oct 5 [cited 2019 Aug 26];215(3):403–10. Available from: http://www.ncbi.nlm.nih.gov/pubmed/2231712

14. Meijenfeldt FAB von, Arkhipova K, Cambuy DD, Coutinho FH, Dutilh BE. Robust taxonomic classification of uncharted microbial sequences and bins with CAT and BAT. bioRxiv [Internet]. 2019 Jan 24 [cited 2019 Aug 26];530188. Available from: https://www.biorxiv.org/content/10.1101/530188v1

15. Ondov BD, Bergman NH, Phillippy AM. Interactive metagenomic visualization in a Web browser. BMC Bioinformatics [Internet]. 2011 Dec 30 [cited 2019 Aug 26];12(1):385. Available from: https://bmcbioinformatics.biomedcentral.com/articles/10.1186/1471-2105-12-385

16. Guo J, Bolduc B, Zayed AA, Varsani A, Dominguez-Huerta G, Delmont TO, et al. VirSorter2: a multi-classifier, expert-guided approach to detect diverse DNA and RNA viruses. Microbiome [Internet]. 2021 Dec 1 [cited 2021 Feb 3];9(1):37. Available from: https://microbiomejournal.biomedcentral.com/articles/10.1186/s40168-020-00990-y

17. Tisza MJ, Belford AK, Domínguez-Huerta G, Bolduc B, Buck CB. Cenote-Taker 2 democratizes virus discovery and sequence annotation. Virus Evol [Internet]. 2021 Jan 20 [cited 2021 Jan 30];7(1). Available from: https://academic.oup.com/ve/article/doi/10.1093/ve/veaa100/6055568

18. Turner D, Kropinski AM, Adriaenssens EM. A Roadmap for Genome-Based Phage Taxonomy. Viruses [Internet]. 2021 Mar 18 [cited 2021 Mar 22];13(3):506. Available from: https://www.mdpi.com/1999-4915/13/3/506

19. Tolstoy I, Kropinski AM, Brister JR. Bacteriophage Taxonomy: An Evolving

Discipline. Methods Mol Biol [Internet]. 2018 [cited 2022 Jan 7];1693:57–71. Available from: https://link.springer.com/protocol/10.1007/978-1-4939-7395-8_6

20. Coutinho FH, Zaragoza-Solas A, López-Pérez M, Barylski J, Zielezinski A, Dutilh BE, et al. RaFAH: Host prediction for viruses of Bacteria and Archaea based on protein content. Patterns. 2021 Jul 9;2(7):100274.

21. Vieira-Silva S, Falony G, Belda E, Nielsen T, Aron-Wisnewsky J, Chakaroun R, et al. Statin therapy is associated with lower prevalence of gut microbiota dysbiosis. Nat 2020 5817808 [Internet]. 2020 May 6 [cited 2022 Jan 7];581(7808):310–5. Available from: https://www.nature.com/articles/s41586-020-2269-x

22. Valles-Colomer M, Bacigalupe R, Vieira-Silva S, Suzuki S, Darzi Y, Tito RY, et al. Variation and transmission of the human gut microbiota across multiple familial generations. Nat Microbiol [Internet]. 2022 Jan 30 [cited 2022 Jan 7];7(1):87–96. Available from: https://www.nature.com/articles/s41564-021-01021-8

23. Shkoporov AN, Clooney AG, Sutton TDS, Ryan FJ, Daly KM, Nolan JA, et al. The Human Gut Virome Is Highly Diverse, Stable, and Individual Specific. Cell Host Microbe [Internet]. 2019 Oct 9 [cited 2019 Nov 14];26(4):527-541.e5. Available from: https://www.sciencedirect.com/science/article/abs/pii/S1931312819304767

24. Nayfach S, Páez-Espino D, Call L, Low SJ, Sberro H, Ivanova NN, et al. Metagenomic compendium of 189,680 DNA viruses from the human gut microbiome. Nat Microbiol [Internet]. 2021 Jul 24 [cited 2021 Jul 1];6(7):960–70. Available from: http://www.nature.com/articles/s41564-021-00928-6

25. Holmes I, Harris K, Quince C. Dirichlet Multinomial Mixtures: Generative Models for Microbial Metagenomics. PLoS One [Internet]. 2012 Feb 3 [cited 2022 Jan

7];7(2):e30126. Available from: https://journals.plos.org/plosone/article?id=10.1371/journal.pone.0030126

26. McMurdie PJ, Holmes S. phyloseq: An R Package for Reproducible Interactive Analysis and Graphics of Microbiome Census Data. PLoS One [Internet]. 2013 Apr 22 [cited 2022 Jan 9];8(4):e61217. Available from: https://journals.plos.org/plosone/article?id=10.1371/journal.pone.0061217

27. Segata N, Izard J, Waldron L, Gevers D, Miropolsky L, Garrett WS, et al. Metagenomic biomarker discovery and explanation. Genome Biol [Internet]. 2011 Jun 24 [cited 2022 Jan 9];12(6):1–18. Available from: https://genomebiology.biomedcentral.com/articles/10.1186/gb-2011-12-6-r60

28. Caenepeel C, Falony G, Machiels K, Verstockt B, Ferrante M, Sabino J, et al. Dysbiosis-associated stool features improve prediction of response to biological therapy (anti-TNF, alpha, anti-integrin and anti-interleukin 12/23) in inflammatory bowel disease. Under Revis Gastroenterol. 2022;

29. Dixon P. VEGAN, a package of R functions for community ecology. Journal of Vegetation Science. 2003.

30. Fong Y, Huang Y, Lemos MP, Juliana McElrath M. Rank-based two-sample tests for paired data with missing values. Biostatistics [Internet]. 2018 Jul 1 [cited 2022 Jan 9];19(3):281–94. Available from: https://academic.oup.com/biostatistics/article/19/3/281/4093659

31. R Development Core Team R. R: A Language and Environment for Statistical Computing. R Foundation for Statistical Computing. 2011.
